# Assessing evidence of immune imprinting in serological patterns of influenza immune response

**DOI:** 10.1101/2025.10.28.25338971

**Authors:** Bernardo García-Carreras, Bingyi Yang, James A. Hay, C. Jessica E. Metcalf, Matt D. T. Hitchings, Claire P. Smith, Siyu Chen, Jonathan M. Read, Huachen Zhu, Chaoqiang Jiang, Kin On Kwok, Steven Riley, Sarah Cobey, Justin Lessler, Derek A. T. Cummings

**Author notes:** **For correspondence:** (BGC); (DATC).

## Abstract

The subtype of the first influenza infection shapes infection risk, illness and mortality associated with subsequent infection subtypes, as well as vaccine efficacy. This phenomenon, termed ‘immune imprinting’, may result from immune recognition rooted in structural similarity of the hemagglutinin protein within subtypes of influenza viruses. However, it is difficult to isolate from cohort effects resulting from the annual dominance of specific influenza subtypes. We used a long-term cohort study in southern China to examine whether hemagglutination inhibition responses show evidence of imprinting, and explore whether such patterns could emerge in the absence of imprinting using simulations. Our statistical analysis reveals patterns consistent with immune imprinting. However, simulations showed that similar patterns can emerge without an imprinting mechanism, meaning these may partly be due to age-related variations in immune responses driven by other factors. This work lays the foundations for further research into immune imprinting while accounting for cohort effects.

## Introduction

The burden of influenza on public health is compounded by the limited efficacy of infiuenza vaccines. Vaccine efficacy is low in part because of barriers posed by pre-existing immunity resulting from prior vaccination or infection. Pre-existing immunity can blunt or amplify responses based on the correspondence of antigens in vaccines with prior exposures (***Reber et al., 2015***; ***Moritzky et al., 2023***). Annual cycles of different influenza can therefore also lead to cohort effects. Immune imprinting represents a specific example of a cohort effect, wherein exposure to specific infiuenza strains during childhood influences the immune system’s memory, impacting future responses to strains based on their similarity to those of early exposures. This phenomenon has been shown to drive age-specific patterns of illness and death, as evidenced by H3N2 underlying a greater proportion of reported cases in older individuals than H1N1 (***Oidtman et al., 2021***), and by models fit to data (***Gostic et al., 2016***; ***Gagnon et al., 2018***; ***Gostic et al., 2019***; ***Arevalo et al., 2020b***). Immune imprinting may pose a substantial challenge to vaccination strategies (***Reber et al., 2015***; ***Moritzky et al., 2023***), as the immune system’s “memory” may either enhance or hinder the effectiveness of subsequent influenza vaccines, depending on the degree of similarity between the circulating strains and those encountered earlier in life. Understanding the mechanisms of immune imprinting is therefore crucial for refining vaccination approaches and addressing the complex dynamics of pre-existing immunity.

A suggested mechanism for imprinting is that the similarity of the hemagglutinin (HA) protein within groups of influenza viruses drives correlated serological responses across subtypes. HA subtypes are clustered into two structural groups, with H1, H2, and H5 among others, in one group, and H3, H7, and H9, among others, in another (***Gostic et al., 2016***). The imprinting hypothesis posits that early exposure to a virus from one group will enhance responses later in life to viruses from the same group relative to viruses of the other group.

One important and straightforward line of evidence for evaluating how imprinting shapes protection against different subtypes is the cross-subtype serological profile of individuals with imprinting resulting from different subtypes. Antibody titers as measured by the hemagglutination inhibition (HI) assay have been shown to correlate with protection (***Hobson et al., 1972***), and have previously been used in studies on antigenic seniority (***Lessler et al., 2012***). To date, however, serological investigations of imprinting have been limited. ***Ranjeva et al. (2019***) fit an individual-level model to longitudinal serological data. They found limited evidence of imprinting, where imprinting was defined as inferred protection from infection, rather than differences in immune response. ***Edler et al. (2024***) observed biases in immune responses to influenza B viruses, with titers tending to be higher against viruses from the same lineage as those encountered early in life. Other studies explored related questions without explicitly testing the immune imprinting hypothesis. ***Miller et al. (2013***), for instance, followed 40 participants over 20 years and provided additional support for antigenic seniority, also reported by ***Lessler et al. (2012***) using a different cohort. However, all participants in the Miller *et al*. study were born prior to 1953, and thus were likely exclusively exposed to one subtype (H1N1) early in life, so that they could not directly test the immune imprinting hypothesis.

Here, we examine immune profiles from a long-term cohort study in southern China to determine if patterns of hemagglutination inhibition responses show evidence of imprinting between subtypes. Specifically, we test the hypothesis that imprinting by a virus from one group (e.g., H1N1 or H2N2) leads to systematically higher immune responses to viruses from the same group, compared to those imprinted by a virus from another group (i.e., H3N2).

## Results

We ask whether an individual’s first influenza subtype exposure (their “imprinting subtype”) leaves a detectable signature in their antibody profile later in life. To investigate this, we used serological data from the Fluscape cohort, a long-term community-based study in southern China that enrolled 2,008 individuals aged 2–95 between 2009 and 2015. For each participant, hemagglutination inhibition (HI) titers were measured against a panel of 21 A(H3N2) strains that circulated between 1968 and 2014, as well as three A(H1N1) strains. Because H3N2 belongs to HA Group 2 while H1N1 and H2N2 both belong to HA Group 1, the imprinting hypothesis makes a specific directional prediction: participants whose earliest influenza exposure was to an H1N1 or H2N2 virus (Group 1) should have systematically lower HI titers against H3N2 strains than participants first exposed to H3N2 (Group 2), and vice versa for H1N1 titers. The central challenge is that imprinting subtype and age are not independent: because specific subtypes dominated circulation during specific historical windows, a participant’s birth year simultaneously determines their likely imprinting subtype and their age at the time of sampling and at the time each test strain was circulating (Fig 1). These age-related quantities are known to independently shape HI titer levels: age at sampling captures the cumulative and waning dynamics of the antibody response over a lifetime, while age at isolation captures the effect whereby individuals tend to mount stronger responses to strains that circulated when they were young. Any statistical approach that controls for age therefore risks absorbing part of the very signal it is trying to detect.

**Figure 1.**
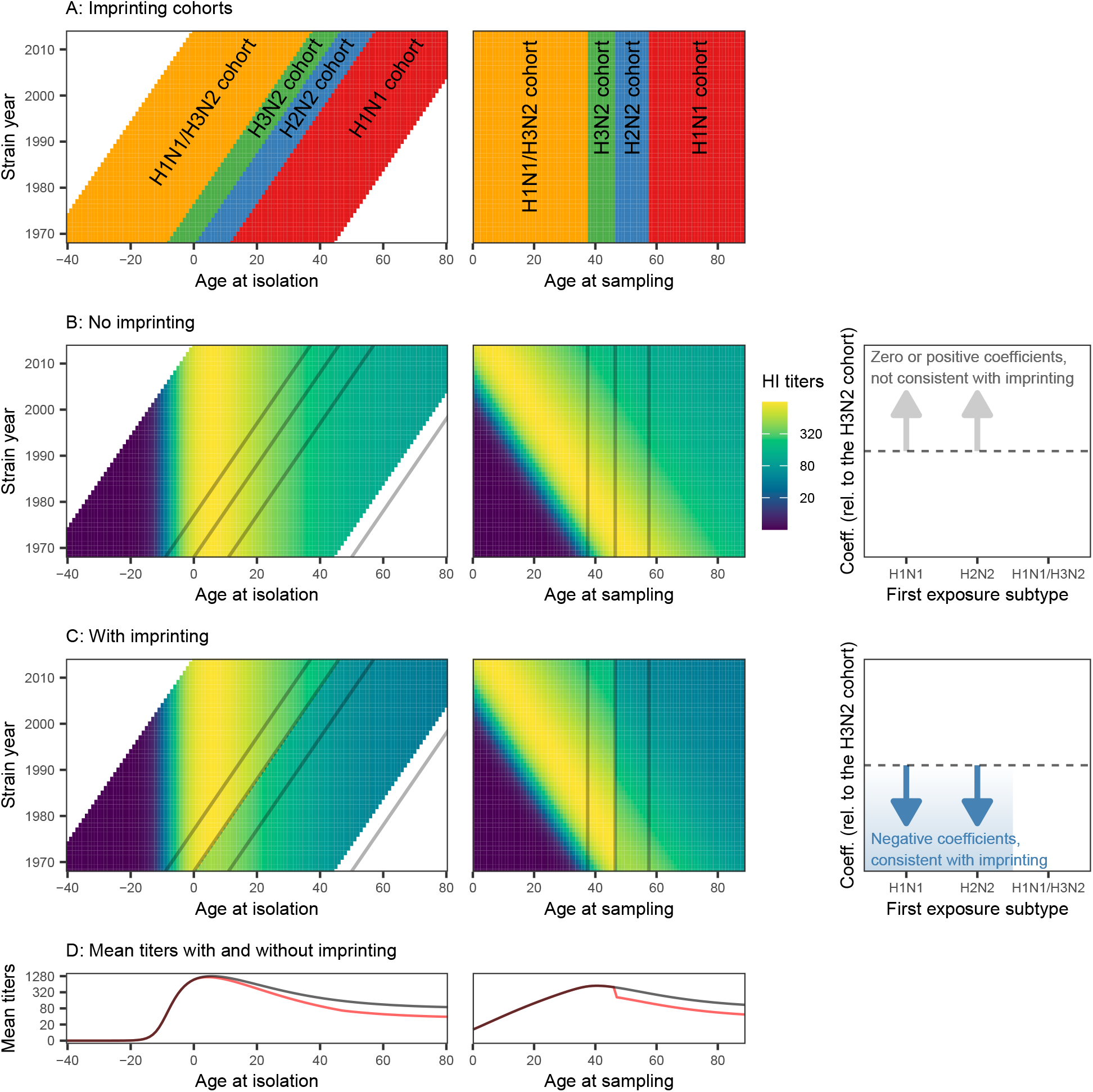
Schematic representation of immune imprinting in our models. (A) Cohorts defined by the subtype of first exposure (assuming infection occurs during the first year of life), as a function of the year a strain was circulating (1968–2014) on the y-axis, and participant age at isolation (the participant’s age when a given strain was circulating) and age at sampling (the participant’s age when the sample was collected, here assumed to be 2014) on the x-axes. (B and C) Idealised H3N2 HI titers as a function of age at isolation and age at sampling. Each coloured tile shows the mean titer for participants of that age (at isolation or at sampling) against the corresponding strain (see Fig 4 for an equivalent heatmap using data). The lines indicate the contours delineating the cohorts shown in (A). In (B), we assume no imprinting, which would yield coefficients for the H1N1 and H2N2 cohorts (relative to the H3N2 cohort reference level) that are either not significantly different from zero, or positive. (C) Same as (B), but with titers for the H1N1 and H2N2 cohorts suppressed to illustrate the hypothesised effect of imprinting. In this case, we would expect significantly negative coefficients for the H1N1 and H2N2 cohorts (i.e., systematically lower titers in the H1N1 and H2N2 cohorts, relative to the H3N2 cohort). (D) Mean titers as a function of age at isolation and age at sampling, without imprinting (black lines), and with the assumed imprinting effect (red lines). Note that these are schematic idealisations, and do not necessarily represent the real patterns in the data.

We fit a suite of generalised additive models (GAMs) to log-transformed HI titers, and varied how age was accounted for: models were fit either without any age adjustment, with a smooth spline on age at sampling only, with a spline on age at isolation only, or with both splines simultaneously (additively). Then, each of these age-adjusted models was fit twice, once without a categorical imprinting variable, and once including it, allowing us to assess both whether imprinting-consistent patterns emerge, and whether including the imprinting variable meaningfully improves model fit as judged by BIC. The imprinting variable encodes each participant’s likely subtype of first exposure (Fig 1). Because that first exposure cannot be observed directly, we used two approaches: a “non-probabilistic” version that assigns a subtype deterministically from birth year (assuming infection always happens within the first year of life), and a “probabilistic” version that draws subtype assignments from empirically estimated probabilities (***Gostic et al., 2016***), repeating this 1000 times to propagate the resulting uncertainty through all model estimates. In both cases, participants born after 1977 (when H1N1 re-emerged alongside H3N2) form a separate mixed category (“H1N1/H3N2”) that is retained in the model but is not the primary focus.

The results of each model are summarised through the estimated coefficients for the imprinting variable, which quantify how much the mean log-transformed HI titers for the H1N1- and H2N2-imprinted cohorts differ from those of the H3N2-imprinted reference cohort, after accounting for all other terms in the model (Fig 1). A coefficient that is significantly negative for the H1N1 and H2N2 cohorts in the H3N2 titer analyses, meaning those cohorts have systematically lower H3N2 titers, would be consistent with the imprinting hypothesis.

We complement the empirical analysis with fits of the same GAM suite of models to output from a mechanistic simulation model (***Yang et al., 2022***) that simulates the evolution of immune responses to H3N2 viruses from 1968 to 2014 for 777 participants, serving as a null model against which to test the imprinting hypothesis. This model does not explicitly include any imprinting mechanism (or any other between-subtype interaction). As a positive control, we also simulated a hypothesised imprinting effect by boosting responses by 50% for participants born on or after 1968, relative to those born earlier (see Methods and Materials).

### We find some support for the imprinting hypothesis

The best model by BIC included age at sampling, age at isolation, and the probabilistic imprinting variable, but only the coefficient for the H2N2 cohort was consistent with the imprinting hypothesis (orange points in Fig 2A and Table S3 in Supplemental Information (SI)). However, this support was sensitive to the choice of age variables and imprinting variables included (Fig 2). We therefore consider our results to weakly support the imprinting hypothesis.

**Figure 2.**
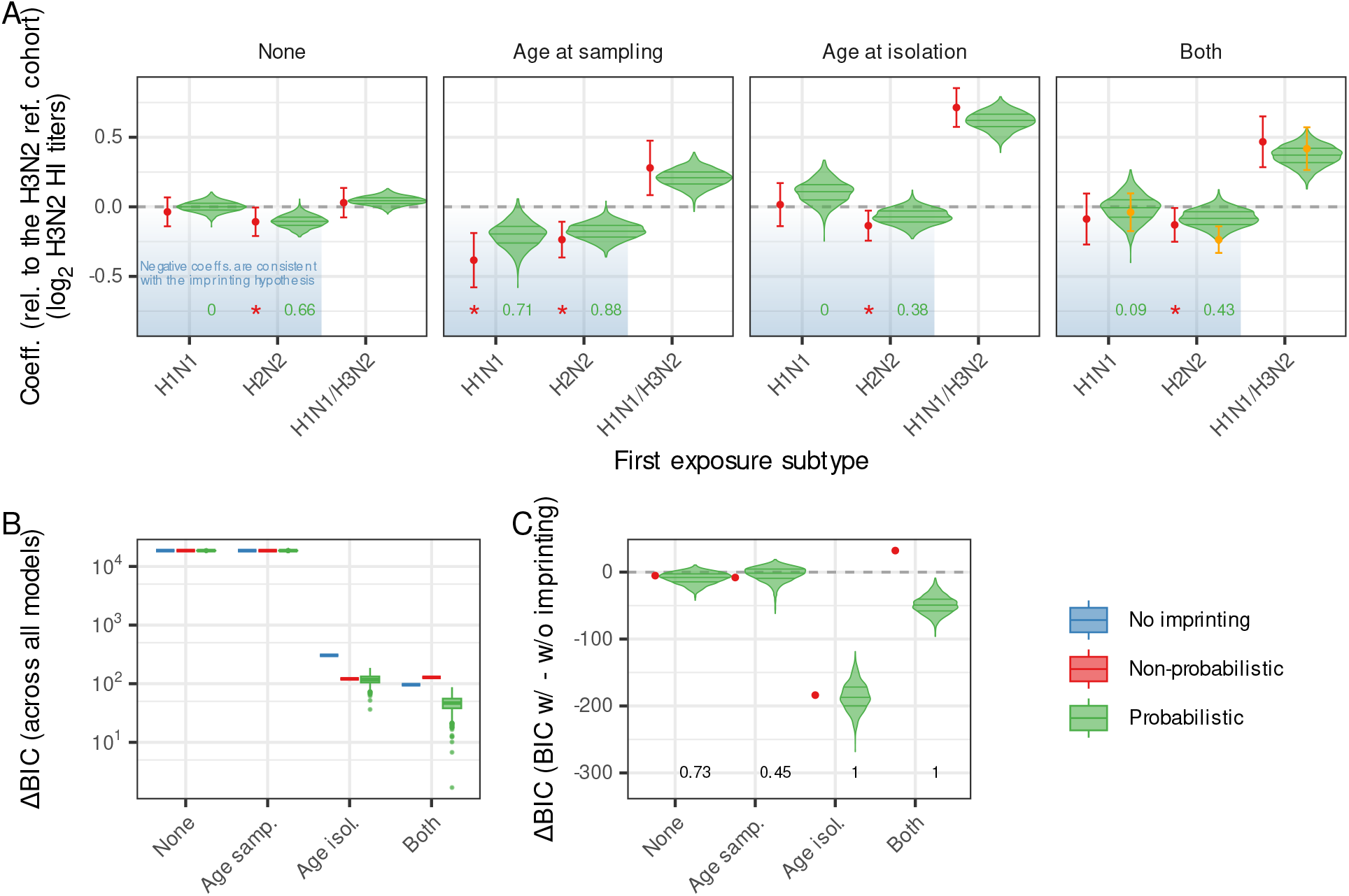
Coefficients for imprinting variables and model comparisons for GAMs fit to H3N2 HI titers. (A) The coefficients for the imprinting variables, relative to the reference H3N2 cohort. Negative coefficients for the H1N1 and H2N2 cohorts (i.e., lower titers relative to the H3N2 cohort; shaded in blue) would be consistent with the imprinting hypothesis. Each panel in (A) represents different adjustments for age, with “Both” indicating models incorporating age at both sampling and isolation. Within each panel, the red points and lines show the coefficients and uncertainty intervals for the non-probabilistic imprinting variable. Red stars highlight coefficients that are statistically consistent with the imprinting hypothesis. The violin plots show the distributions of 1000 coefficients for the probabilistic imprinting variable, with numbers below indicating the proportion of the coefficients that are statistically significant and consistent with the imprinting hypothesis. The orange points and lines show the coefficients for the best model by BIC. See Table S3 in SI for the coefficients for the three best models by BIC. (B) BIC values relative to the model with the lowest BIC, across all models, both without and with imprinting variables (non-probabilistic and probabilistic). (C) The differences in BICs between models with and without the imprinting variable; negative values suggest better performance of the model with the imprinting variable. The numbers below the violin plots in (C) show the proportion of comparisons for which the BIC difference is three or more in favour of the model with the imprinting variable. See Fig S11 in SI for analogous results using deviance explained and root mean square error (RMSE) rather than BIC.

More generally, when using the non-probabilistic imprinting variable, coefficients aligned with the imprinting hypothesis when the model included age at sampling (Fig 2A). Conversely, when age was not included, the coefficients were not significant; when only age at isolation was included, only the coefficient for the H2N2 cohort was significant. In contrast, for the probabilistic imprinting variable, only models incorporating age at sampling yielded coefficients largely in line with the imprinting hypothesis. However, when age was not included, or when only age at isolation was included, coefficients for the H1N1 cohort (and to a lesser extent the H2N2 cohort) did not significantly deviate from those for the H3N2 cohort. Models that included age at isolation were significantly better (by BIC) compared to models that only included age at sampling (Fig 2B). The inclusion of both age variables did not consistently improve model performance relative to including only age at isolation. Only when age at isolation was included did models with the imprinting variables consistently outperform models without it (Fig 2C). However, results for models including both age splines should be interpreted with caution, given that the inclusion of an imprinting variable led to substantial changes in the other splines and coefficients included in the models (see Fig S15 in contrast to Figs S12–S14 in SI).

When fitting to H1N1 titers, the best models included the probabilistic imprinting variable. Unlike the H3N2 results, the best model by BIC included age at sampling (Fig S16 and Table S5 in SI). None of the coefficients in these models were consistent with the imprinting hypothesis. More broadly, when age at isolation was included, coefficients for the H1N1 cohort were higher than those for the H3N2 cohort, aligning with the imprinting hypothesis (Fig S16 in SI). However, the coefficients for the H2N2 cohort did not clearly deviate from those for the H3N2 cohort.

### Simulation models without imprinting can produce similar results

As in the models fit to the H3N2 titer data, the best model fit by BIC to the null simulation model (without an imprinting effect) included age at isolation and the probabilistic imprinting variable, although, unlike the fits to the H3N2 titer data, the coefficients did not support the imprinting hypothesis.

More broadly, however, there are clear similarities between the coefficient patterns observed in fits to the data, and those for from the null model, which lacks any imprinting mechanism (Fig 3A). Models incorporating both age variables provide minimal support for the imprinting hypothesis, while models including only age at isolation suggest that only the H2N2 cohort has systematically lower titers than the H3N2 cohort. Nonetheless, inclusion of the imprinting variable in the models is still strongly favoured (by BIC; Fig 3C). Including age at sampling in a model already incorporating age at isolation does not significantly improve model fit (by BIC; Fig 3B,C).

**Figure 3.**
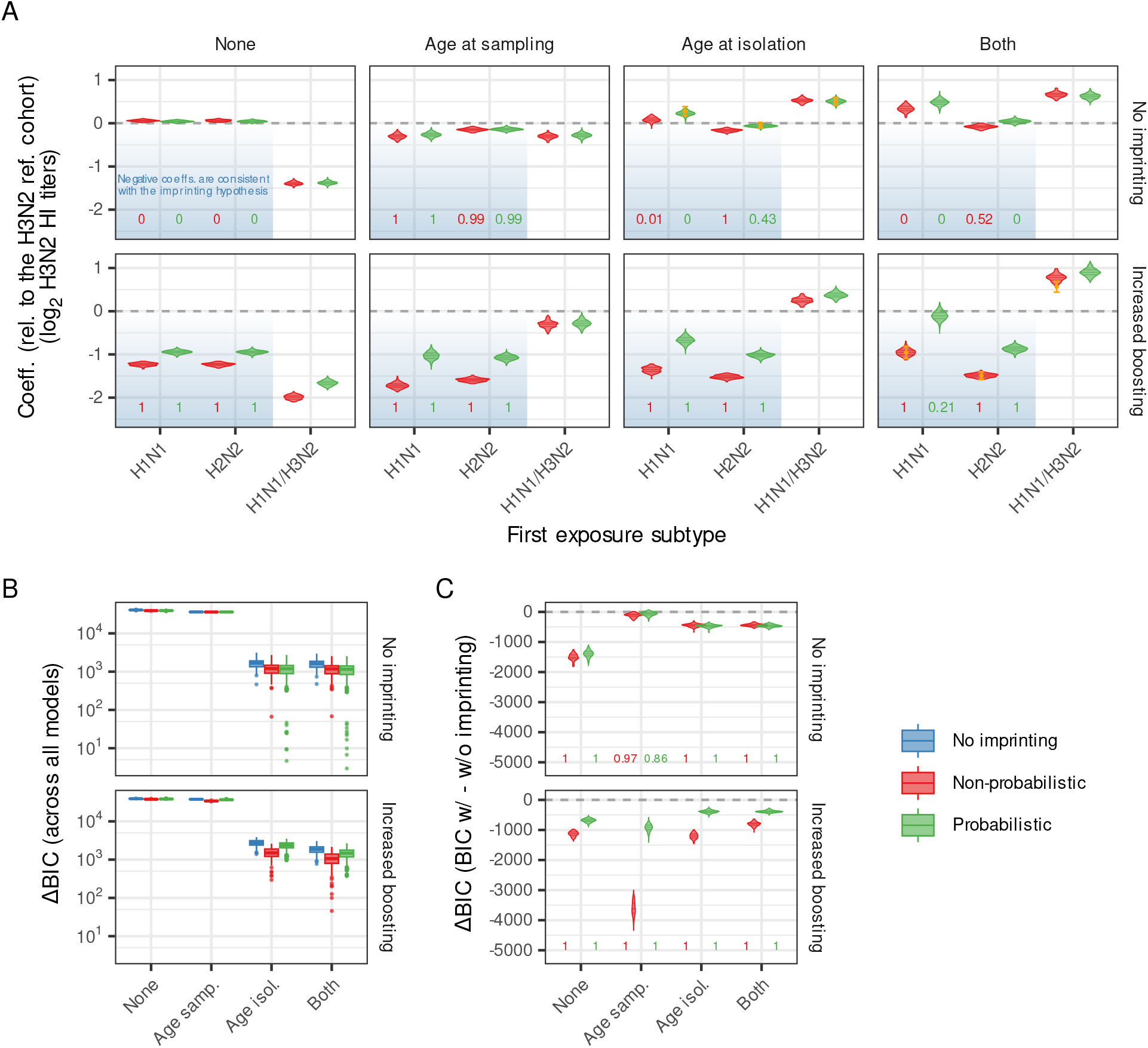
Coefficients for imprinting variables and model performance comparisons for GAMs fit to simulated H3N2 HI titers. The coefficients are shown for fits to both the null model (without an imprinting mechanism) and the positive control model (including imprinting via increased boosting; Methods and Materials). (A) The coefficients for the imprinting variables, relative to the reference H3N2 cohort. Negative coefficients for the H1N1 and H2N2 cohorts (i.e., lower titers relative to the H3N2 cohort; shaded in blue) would be consistent with the imprinting hypothesis. Each panel in (A) represents different adjustments for age, with “Both” indicating models incorporating age at both sampling and at isolation. Within each panel, the red violin plots show the distributions of the coefficients for the non-probabilistic imprinting variable (one for each of the 100 stochastic realisations of the model). The green violin plots show the distributions of 1000 coefficients for the probabilistic imprinting variable. The numbers below the violin plots indicate the proportion of coefficients that are statistically significant and consistent with the imprinting hypothesis (i.e., significantly lower than those for the reference H3N2 cohort). Orange points and lines indicate the coefficients and 95% confidence intervals for the best models by BIC. (B) BIC values are relative to the model with the lowest BIC, across all models, both without and with imprinting variables (non-probabilistic and probabilistic). (C) The differences in BICs between models with and without the imprinting variable; negative values indicate better performance of the model with the imprinting variable. The numbers below the violin plots in (C) show the proportions of the comparisons for which the difference in BIC is three or more in favour of the model with the imprinting variable.

When the effects of an imprinting mechanism are included in the model (the positive control model), the imprinting variable successfully identifies the pattern, producing coefficients clearly consistent with the imprinting hypothesis (Fig 3A). The signal is strongest for the non-probabilistic imprinting variable (as expected, given that the imprinting mechanism in the model is encoded in the same way). However, when both age variables are included, the signal for the H1N1 cohort is diminished for the probabilistic imprinting variable, perhaps explaining why, in our analyses of the data, coefficients for the H2N2 cohort were more consistently in line with the imprinting hypothesis than those for the H1N1 cohort.

## Discussion

While a wide range of data on disease and mortality indicates evidence for immune imprinting, our analysis demonstrates that patterns in serological data are somewhat equivocal. Statistical models of our cohort HI titers showed evidence of immune imprinting; however, this support was dependent on the other variables included in the model and significance patterns were also sensitive to the specific models considered. Evidence was also stronger for H2N2 imprinting being associated with reduced H3N2 titers than for other combinations (a pattern corroborated even in our positive control simulation). Statistical patterns consistent with immune imprinting (i.e., systematically lower HI titers against H3N2 strains in cohorts likely first exposed to and infected by H1N1 and H2N2) could be reproduced by a simulation model lacking an explicit imprinting mechanism or subtype-subtype interactions, although the best-fitting models had coefficients inconsistent with the imprinting hypothesis.

Nevertheless, these results do not represent evidence for the absence of immune imprinting. Indeed, there are many factors that may explain why we might not detect a clear imprinting effect in our analysis of serology, even if the mechanism were at play (***Oidtman et al., 2021***). HI assays quantify titers of antibodies that predominantly bind to sites on the HA head (***Miller et al., 2013***), while imprinting may be mainly driven by antibody responses to the HA stalk (***Gostic et al., 2016***; ***Knight et al., 2020***; ***Arevalo et al., 2020a***), by other components of immune memory (***Nelson and Sant, 2019***), or by the quantity or specificity of antibodies (***Oidtman et al., 2021***). Stalk-specific antibodies produced in response to an imprinting subtype have been shown to weakly bind to virus strains of a different subtype (***Arevalo et al., 2020a***). This could lead to more infections, and consequently, a relative increase in head-specific antibodies and higher HI titers against strains of the non-imprinting subtype. Furthermore, prior evidence for imprinting relied on reported severe and fatal cases, suggesting that imprinting may primarily reduce disease severity. However, the relationship between HI titers, protection, and disease severity is complex, and likely a function of age, subtypes, and other factors (***Fox et al., 2015***; ***Ranjeva et al., 2019***; ***Oidtman et al., 2021***). Our dataset may include participants who were either uninfected or asymptomatic; in these cases, immune imprinting could be absent or its effects more subtle. Immune imprinting might also not be solely mediated by antibody responses; other branches of the immune response (e.g., CD4 T cells; ***Nelson and Sant (2019***)) may play a role, although previous studies indicating stratification in immune responses by hemagglutinin type suggest that these effects would still be subtype-specific (***Gostic et al., 2016, 2019***).

In ascribing a subtype of likely first exposure to participants, we made assumptions about how early children are likely to be infected. The assumptions ranged from a first infection being certain within the first year, to possible within the first twelve years (assumption made by ***Gostic et al. (2019***)). The reality likely lies between the two. ***Brouwer et al. (2022***) found that a significant proportion of children under one year of age have antibodies against influenza strains, but this fraction decreased with the number of years since a new antigenic cluster was introduced. Furthermore, whether imprinting is driven exclusively by the first exposure, or more generally by the first few exposures, also remains an open question (***Oidtman et al., 2021***) of particular significance in understanding the role immune imprinting may be playing post-1977, due to the co-circulation of H1N1 and H3N2.

Given our focal question, accounting for participant infection history was necessary in all our analyses, which we achieved by including participant age (at sampling, at isolation, or both) in the model. However, any imprinting effect would manifest as a birth cohort effect. To account for age effects, we used GAMs, a highly flexible non-parametric approach, and then included a categorical variable to detect systematic differences between birth cohorts consistent with the imprinting hypothesis. The implicit presumption is that systematic patterns caused by imprinting would be more parsimoniously captured by this categorical variable rather than in the age splines. To minimise the risk of the GAMs masking patterns attributable to imprinting, we performed model selection with double penalisation within the GAMs. Despite these efforts, the splines and other model coefficients may still have absorbed some of the imprinting signal (if there was one). For instance, coefficients for the H1N1/H3N2 cohort were often significantly positive, likely because this cohort comprises younger participants more likely to have had a recent infection. These systematically higher titers were to some extent captured by both the imprinting variable and the age splines. Results from models using both age splines should be interpreted with caution. In these models, most coefficients and splines changed substantially when the imprinting variable was added compared to when it was absent, indicating that any imprinting effect could be masked by an interplay of compensating factors (see Section “Sensitivity analyses” in SI). The compensation in coefficients and splines observed when the imprinting variable is absent also explains why RMSE differences between models with and without imprinting are small (Fig S11 in SI). However, these small differences should not be taken as evidence against an imprinting effect, given our aim is to test the hypothesis, not to demonstrate that the inclusion of the imprinting variable improves prediction. Our approach did detect the imprinting mechanism when its effects were included in the simulation model. Specifically, we simulated the imprinting effects by increasing boosting by 50% for participants imprinted by a virus of the same group as the infecting strain. Were the effects of imprinting more subtle, or operating through other mechanisms, the resulting coefficients might be harder to distinguish from those of the model without an imprinting mechanism.

As a control for participant infection history in the context of possible cohort effects driven by years of different strains circulating, we fit the same statistical models to the output of a simulation model. This model did not include subtype-subtype interactions and made some simplifying assumptions. For example, the model assumes that antigenic change between strains varies linearly with time. The model was parameterised using a subset of the cohort data analysed here, as well as data from a separate cohort in Vietnam. It is therefore possible that patterns consistent with immune imprinting were implicitly subsumed in the resulting parameterisation. This could account for the observed similarity between the coefficients of the models fit to the simulation output and those fit to the data.

To further explore what patterns would be expected were imprinting to be at play, the simulation model could be refined to introduce immune imprinting by explicitly including other subtypes and their interactions.

A greater understanding of immune responses and the interplay between viral evolution and the life course of immunity to influenza holds significant promise for shaping more effective vaccine policies and formulations. Determining the mechanisms underlying immune imprinting could lead to better predictions of how individuals may respond, based on their prior exposures, to antigenic drift and shift in circulating strains, to influenza vaccine candidates, and to non-human infiuenza viruses. Our analysis demonstrates that imprinting-like patterns can emerge from the confiuence of age and temporal phenomena, even in the absence of the hypothesised immunological mechanisms of imprinting. The unique experiences of specific cohorts, such as the timing of particular infections or time since the introduction of a new antigenic cluster, can result in apparent subtype-subtype interactions (***Brouwer et al., 2022***). This highlights the complexity of interpreting serological patterns in the context of age and temporal dynamics. These findings may also apply to other pathogen systems characterised by complex interactions between host exposure history and rapidly evolving (e.g., SARS-CoV-2) or co-circulating (e.g., dengue) viruses.

## Methods and Materials

### Ethical approval

The following institutional review boards approved the study protocols: Johns Hopkins Bloomberg School of Public Health (IRB 1716), University of Florida (IRB201601953), University of Liverpool, University of Hong Kong (UW 09-020), and Guangzhou No. 12 Hospital (‘Research on human infiuenza virus immunity in Southern China’). Written informed consent was obtained from all participants over 12 years old; verbal assent was obtained from participants 12 years old or younger. Written permission from a legally authorized representative was obtained for all participants under 18 years old.

### Data

The Fluscape cohort study is described by ***Jiang et al. (2016***). In summary, between 2009 and 2015, 4130 serum samples from 2008 individuals aged 2 to 95 years were collected in Guangzhou, China, across 40 locations and 920 households. From these, 61,614 HI titers against 21 A(H3N2) strains that circulated in the region over 47 years (1968–2014, with the exception of X31, a synthetic 1970 strain), and 5845 HI titers against three A(H1N1) strains (1977, 2005, and 2009), were measured. Samples were collected at roughly annual intervals, and on average, two samples were collected per participant. Serum samples were processed and titers estimated in three batches (lab rounds). See Figs 1 and 4 for an overview. Of the cohort, ≈ 75% reported no prior vaccination at all sampling visits, although vaccination reporting was inconsistent.

**Figure 4.**
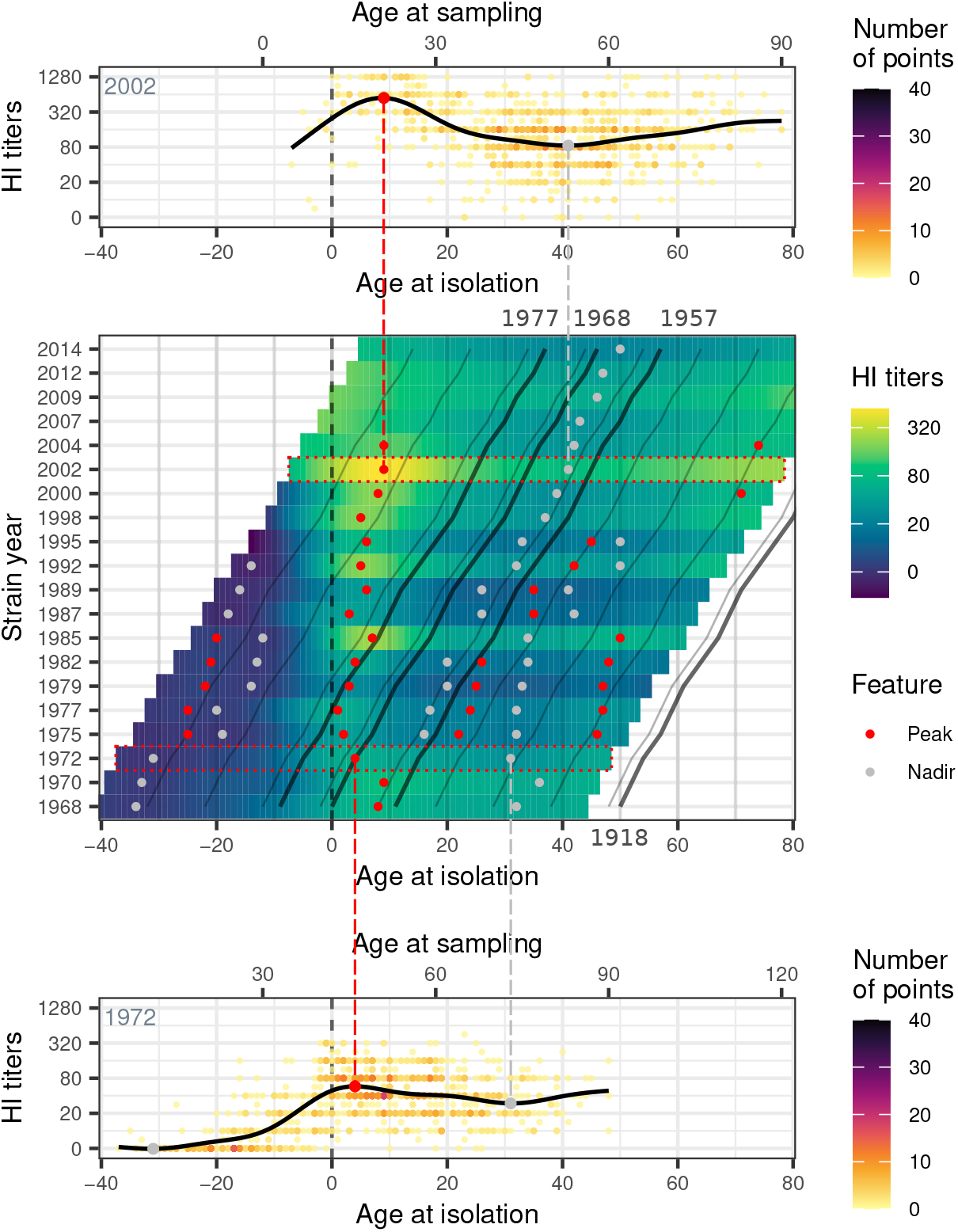
Overview of the Fluscape dataset, with emphasis on the patterns of HI titers with age. The middle panel displays HI titers predicted for each H3N2 strain as a function of age at isolation, for visit 4 (2014–2015) and lab round 3. Predictions were generated using GAMs fitted independently to each strain, including household ID as a random effect (these differ from the GAMs used in the main analysis, in which all strains were included simultaneously). Red and grey points indicate the locations of peaks and nadirs in the predicted HI titers for each strain. The diagonal contour lines follow birth cohorts, with thicker lines representing birth cohorts of specific influenza relevance: 1918, 1957, 1968, 1977. For illustration, we select two strains (1972 on the bottom panel and 2002 on the top panel), both highlighted in the heatmap with red dotted lines, and overlay the predicted titers from the GAM on the data used for fitting. The colour of the points in the top and bottom panels indicates the number of overlaid points. Note the systematically higher titers in individuals who were young when each strain circulated (as previously observed by ***Lessler et al. (2012***) using a subset of the same data), and the systematically low titers in individuals born shortly after the introduction of H2N2 in 1957. In an analogous heatmap expressed as a function of age at sampling, the left and right edges of each row would vertically line up.

### Statistical approach

We fit competing generalised additive models (GAMs) using the ‘bam’ function in the ‘mgcv’ package in R (***R Development Core Team, 2023***; ***Wood, 2011***). We transformed titers (expressed as the inverse of the two-fold serial dilutions) using *z* = log_2_(*x*/5), where *x* is the titer, setting *z* = 0 when titers were undetectable. The log titers (*z*) were the dependent variable in the models. These models assume normally distributed residuals.

Immune responses are known to vary systematically with age (see Fig 4; ***Lessler et al. (2012***); ***Yang et al. (2020***)), and we control for age using two variables: the age of the participant at the time of sampling (age at sampling), and the age the participant would have been when a given strain was circulating (age at isolation; Fig 1).

We fit a suite of models, accounting for age in different ways: (i) a model without age; (ii) a model with a thin plate regression spline on age at sampling; (iii) a model with a spline on age at isolation; and (iv) a model with both, one spline on age at sampling and the other on age at isolation (additively). In all models, we additionally account for visit number (as a categorical variable), time since the start of the study (with a spline), and participant gender. Finally, preliminary analyses showed that titers vary systematically across lab rounds and strains (e.g., see Fig 4). We therefore include an interaction between strain and lab round (and the main effect of each). To account for pseudoreplication (within-person correlations), we include a random effect for participant ID (encoded using bs = “re” in ‘mgcv’). See Table S1 in SI for the rationale behind each parameter. To mitigate overfitting, we use double penalisation (via the select = TRUE argument in the ‘mgcv’ R package). We use restricted maximum likelihood to select the smoothing parameters.

Each of these models was fit with and without an “imprinting” variable, which is the variable of interest. It encodes the likely subtype of first exposure based on year of birth. The coefficients of this variable (one for each subtype) indicate whether, when accounting for the aforementioned factors, participants imprinted with a given subtype tends to have systematically higher or lower H3N2 or H1N1 titers. The hypothesis that imprinting is detectable in serological data would correspond to systematically lower H3N2 titers in participants imprinted with H1N1 or H2N2 compared to those imprinted with H3N2. Conversely, for the analyses of H1N1 titers, the expectation is reversed: participants imprinted with H3N2 should have systematically lower H1N1 titers compared to those imprinted with H1N1 or H2N2.

We use two approaches to determine the subtype of first exposure. In the first approach, we assume that participants are exposed to influenza within their first year of life, so that the subtype of first exposure is determined solely by year of birth (see Fig 5). For example, the subtype of first exposure would be H1N1 for those born before 1957, H2N2 for participants born between 1957 and 1967 (inclusive), and H3N2 for those born between 1968 and 1976. In 1977, H1N1 reemerged, and has since co-circulated with H3N2. We treat this as a separate mixed category (“H1N1/H3N2”), and explain the reason in Section “Rationale for grouping the H1N1 and H3N2 cohorts for participants born after 1977” in SI. We refer to this approach as the “non-probabilistic” immune imprinting variable.

**Figure 5.**
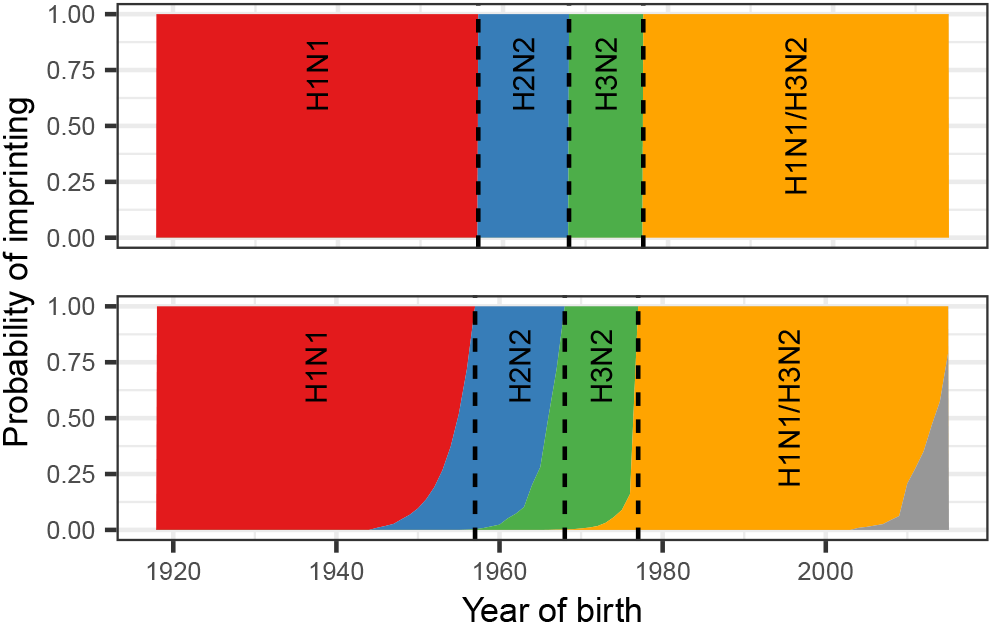
Proportions of each birth cohort with an initial exposure to each subtype. These proportions correspond to 2014 in China, as obtained from ***Gostic et al. (2016***). The top panel assumes that participants get infected in their first year of life, while the bottom panel relaxes this assumption, leading to non-zero probabilities of being infected and imprinted to more than one subtype in the years prior to the introduction of a new subtype (***Gostic et al., 2016***).

In the second approach, we assume that individuals may not be exposed to infiuenza within their first year of life, so that those born prior to the introduction of a new subtype may still have a non-zero probability of being imprinted with it. For example, a participant born in 1955, during the period when H1N1 was circulating, may still have been imprinted with H2N2 (which emerged in 1957). For this purpose, we use the subtype imprinting probabilities as a function of year of birth estimated by ***Gostic et al. (2016***) for China (see Fig 5). For each participant, we sampled the subtype of first exposure from these probabilities. We then fit all models 1000 times, resampling the subtype of first exposure for each participant each time. As in the first approach, we treat the period characterised by the reintroduction of H1N1 as a mixed H1N1/H3N2 category. We refer to this approach as the “probabilistic” immune imprinting variable.

The model formula thus takes the following form:

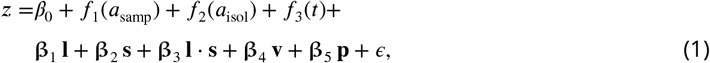

where *f*_1_, *f*_2_, and *f*_3_ are smooth functions, **β**_0_, **β**_1_, … **β**_5_ are vectors of coefficients, *a*_samp_ and *a*_isol_ are the ages at sampling and isolation, respectively, **1** is the lab round, **s** is the strain, **v** is the visit, **p** the imprinting variable, and ϵ the error term. The age terms can both be excluded or included, or only included one at a time. Models are run including and excluding the imprinting variable. See Table S2 in SI for a list of the set of models fit.

### Simulation models

We use previously described simulations (***Yang et al., 2022***) as null models for testing the imprinting hypothesis. This mechanistic model focuses on the interplay between antibody responses, incorporating within-subtype cross-reactivity and antibody-level-dependent infection risk, with no imprinting mechanism (or any other between-subtype interaction) explicitly included. We refer to this model as the “null model”. In brief, the model simulates the evolution of immune responses to A(H3N2) from 1968 to 2014, for a subset of 777 participants with a similar birth year distribution as those in our study. Prior to simulating each season, we calculated each participant’s probability of infection based on their immune profile and immunity-dependent protection (***Hoa et al., 2022***). Infection outcomes were sampled annually for each participant, and immune profiles updated according to these outcomes and cross-reactivity. We assumed that an A(H3N2) virus cross-reacts with viruses isolated within 10 years of its own isolation, consistent with previous estimates of the rate of evolution (***Fonville et al., 2014***; ***Kucharski et al., 2018***). The overall population attack rate in the population was assumed to be constant at 20%.

As a positive control, we also ran simulations mimicking the hypothesised imprinting effect by increasing the antigenic-targeted boosting of participants born on or after 1968 by 50% (on the logscale) relative to those born prior to 1968. We refer to this model as the “positive control model”.

From the simulation output, we extracted the titers from two visits (2004 and 2014) against 21 H3N2 strains circulating in the same years as those in the data.

Output from the models was analysed using the same approach described above (“Statistical approach”). Here, however, we had 100 stochastic realisations of the model, yielding 100 sets of coefficients for the non-probabilistic imprinting variable. For each of the 100 model realisations, we drew 10 samples of the subtype of first exposure when using the probabilistic imprinting variable, producing 1000 sets of coefficients in total. These 1000 coefficients therefore incorporate both uncertainty in the sampling of the imprinting variable (as in the analogous analyses of the data) and uncertainty in the simulation itself through the 100 model realisations.

### Sensitivity analyses

We tested the robustness of our results with several sensitivity analyses. The GAMs we fit assume normally distributed residuals; we also fit negative binomial GAMs and find that qualitative patterns and significance of coefficients do not change. Several of the parameters we fit are correlated with the imprinting variable, raising the question of parameter identifiability, and whether, or how much of, a possible underlying imprinting signal may be absorbed by parameters other than the imprinting variable. To this end, we re-fit the same models to data generated from the GAMs, with and without an artificial imprinting effect. Parameters were largely recovered, although we also show how an imprinting effect may be partly absorbed by parameters other than the imprinting variable. We also assess whether our results change when accounting for maternal antibodies, finding little change in the patterns observed. We also fit our models to subsets of the data. Results show little change in patterns when fitting models to strains that circulated prior to 2005, but coefficients were overall less consistent with the imprinting hypothesis when fitting to participants born prior to 1977. Finally, we check whether our results based on the simulation output are specific to the demographic profile of the participants. Results were unchanged when fitting our models to simulated HI titers for a population with an approximately uniform birth year distribution. All of these analyses, and the corresponding results, are described in further detail in Section “Sensitivity analyses” in SI.

## Data Availability

Data used in the analyses are available at https://github.com/UF-IDD/influenza_imprinting.

