## Supplemental Information for "Assessing evidence of immune imprinting in serological patterns of influenza immune response"

Supplementary text.
Figures S1–S16.
Tables S1–S6.

### Model parameters

Table S1: **Parameters included in the GAM models.**

| Parameter | Explanation | Form | Inclusion |
|---|---|---|---|
| Age at sampling | Young individuals might have systematically higher titers at the time of sampling due to more recent infections. | Spline | Data, simulations |
| Age at isolation | Individuals have highest titers to strains encountered early in life, and are expected to have no antibodies to strains which circulated significantly before they were born. | Spline | Data, simulations |
| Lab round × strain | Titers can vary systematically by strain and lab round (e.g., a strain might elicit higher titers than other strains, and that can vary as a function of the lab round in which the samples were processed). | Categorical (21 strains, 3 lab rounds, total 31 levels) | Data (only strain in simulations) |
| Time since start of study | Titers may vary systematically with time (for instance, after a large H3N2 season). | Spline | Data |
| Visit | Samples were collected in four different visits over time, to account for small variations in study team members and procedures. This term accounts for any small biases that may have been introduced as a result. | Categorical (4 levels) | Data, simulations |
| Imprinting | The hypothesis is that there may be systematic shifts in titers as a function of the subtype of first exposure. This is a birth cohort effect and thus collinear with the age at sampling. The presumption is that if these shifts align with the imprinting groups (H1N1 and H2N2; H3N2), then they would be more parsimoniously captured here. Sets of models are fit with and without this variable. | Categorical (4 levels) | Data, simulations |
| Individual-level effect | Because we have multiple titers per participant, this accounts for within-person correlations. | Random effect | Data, simulations |

Table S2: **Sets of models fit to the data and simulation output.**

| | Age term(s) | Imprinting term | Number of models |
|---|---|---|---|
| 1 | No age | No imprinting | 1 |
| 2 | | Non-probabilistic imprinting | 1 |
| 3 | | Probabilistic imprinting | 1000 |
| 4 | Age at sampling | No imprinting | 1 |
| 5 | | Non-probabilistic imprinting | 1 |
| 6 | | Probabilistic imprinting | 1000 |
| 7 | Age at isolation | No imprinting | 1 |
| 8 | | Non-probabilistic imprinting | 1 |
| 9 | | Probabilistic imprinting | 1000 |
| 10 | Age at sampling + age at isolation | No imprinting | 1 |
| 11 | | Non-probabilistic imprinting | 1 |
| 12 | | Probabilistic imprinting | 1000 |

#### Rationale for grouping participants born after 1977 into a single H1N1/H3N2 cohort

After 1977, H1N1 reemerged and has been co-circulating with H3N2 ever since. In our model, we group individuals who may have been exposed to both subtypes early in life into a combined H1N1/H3N2 cohort. We could have alternatively used the probabilities from Gostic et al. (2016) to randomly assign one of the two subtypes to each participant born after 1977. However, we chose not to do so for two main reasons:

(i) The probabilities of being exposed to each subtype after 1977 are fairly similar, meaning participants would have a relatively even chance of first encountering either subtype. As a result, we would likely need significantly more than the 1000 samples we use in our analysis to get to a "true" allocation of subtypes. Allocating a subtype of first exposure to these participants could also obscure the signal from the pre-1957 H1N1 cohort and the 1968–1977 H3N2 cohort, making it harder to detect any potential patterns present in those earlier groups.

(ii) Whether imprinting is influenced only by a participant's first exposure, or by the first several exposures, is still an open question. If imprinting is driven by multiple exposures, it could mean that participants born after 1977, who may well have been exposed to both subtypes early in life, might experience imprinting differently compared to participants who were likely exclusively exposed to just one subtype early in life.

### Sensitivity analyses

#### Distribution of residuals

The HI titers being modelled are discrete and equal to or greater than zero. However, they represent an underlying continuous process where the true titer values have been rounded to whole numbers by the dilution series used. When considering the underlying continuous nature of the data, the assumption of normally-distributed residuals in the GAMs we use may be reasonable. On the other hand, when viewed as interval-censored data, this assumption may be less adequate. To address this, we performed a robustness analysis by fitting a subset of the models using negative binomial GAMs (with `family = nb()`).

Results in Supplemental Information (SI) Fig S1 show that while the absolute magnitude of the imprinting coefficients for the negative binomial GAMs tended to be smaller, the qualitative pattern and significance of the coefficients closely followed those presented in the main text.

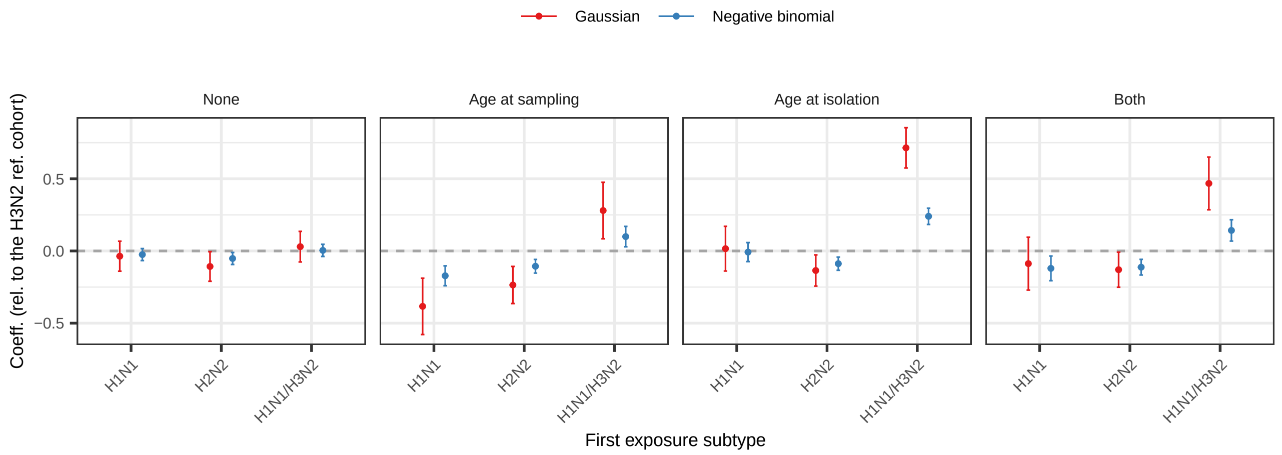

Figure S1: **Coefficients for different assumptions on the distribution of residuals.** Comparison of the models assuming normally distributed residuals (the models used in the main text), and the negative binomial GAMs, for models with the non-probabilistic imprinting variable.

#### Parameter identifiability

The model includes correlated variables, raising the question of parameter identifiability. To test this, we first generated datasets using models fit without the imprinting variable. We then refit the model (without an imprinting variable) to the predicted data (with a small amount of noise added). Next, we introduced an artificial imprinting effect by subtracting 0.05 from the predicted H1N1 and H2N2 cohort titers, and adding 0.05 to the predicted H3N2 cohort titers. Finally, we refit the models with the non-probabilistic imprinting variable. The expectation is to recover the original coefficients, while successfully detecting the imposed imprinting effect (a 0.1 difference in log titer between the H1N1 and H2N2 cohorts, and the H3N2 cohort).

The results, shown in SI Figs S2–S5, confirm that coefficients are recovered when the model is fit to the predicted data, with minor changes to some of the coefficients and splines (compare red and blue points and lines). When the artificial imprinting mechanism is introduced, the model (with the imprinting variable) for the most part correctly detects the effect (panels A). Our results, however, also illustrate the potential for parameters other than the imprinting variable to absorb birth cohort effects. There were minor variations in strain coefficients, particularly for strains of the mid-70s and 80s for lab rounds 1 and 2. More significantly, the inclusion of an artificial imprinting effect and variable led to a downward shift in the age at sampling spline for younger individuals, and an upward shift for older individuals (SI Figs S3D and S5D). These shifts happen despite also detecting systematic differences in titers in the imprinting variable, meaning that some of the signal is absorbed in the age at sampling spline. On the other hand, the inclusion of age at isolation in the model led to higher than expected coefficients for the H1N1/H3N2 cohort (SI Figs S4A and S5A). At the same time, the shape of the spline for time since the start of the study was also affected by the inclusion of age at isolation (compare panels C in SI Figs S2–S5), an effect compensated by changes in the coefficients for the visits (not shown). When both age variables are included, however, the introduction of an imprinting effect leads to significant changes in the strain and lab round coefficients, and in the age splines (SI Fig S5).

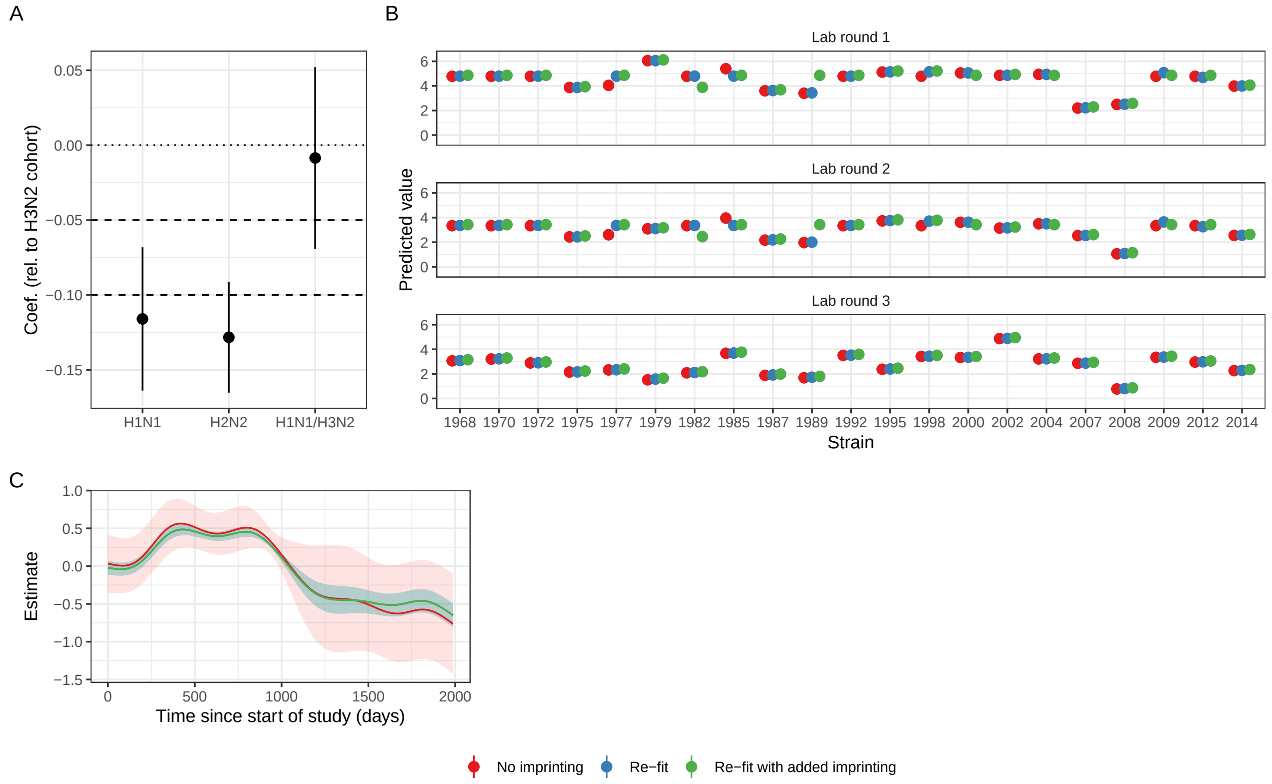

Figure S2: **Comparison of the coefficients and splines to check for parameter identifiability.** We first fit a model without age variables or imprinting ("no imprinting"), generate a dataset with the model, add an artificial imprinting effect, and refit the same model with the addition of the non-probabilistic imprinting variable ("re-fit with imprinting"). We expect the imprinting variable coefficients to be −0.1 for the H1N1 and H2N2 cohorts and −0.5 for the H1N1/H3N2 cohorts (dashed lines in panel A), and otherwise expect all other coefficients to be the same.

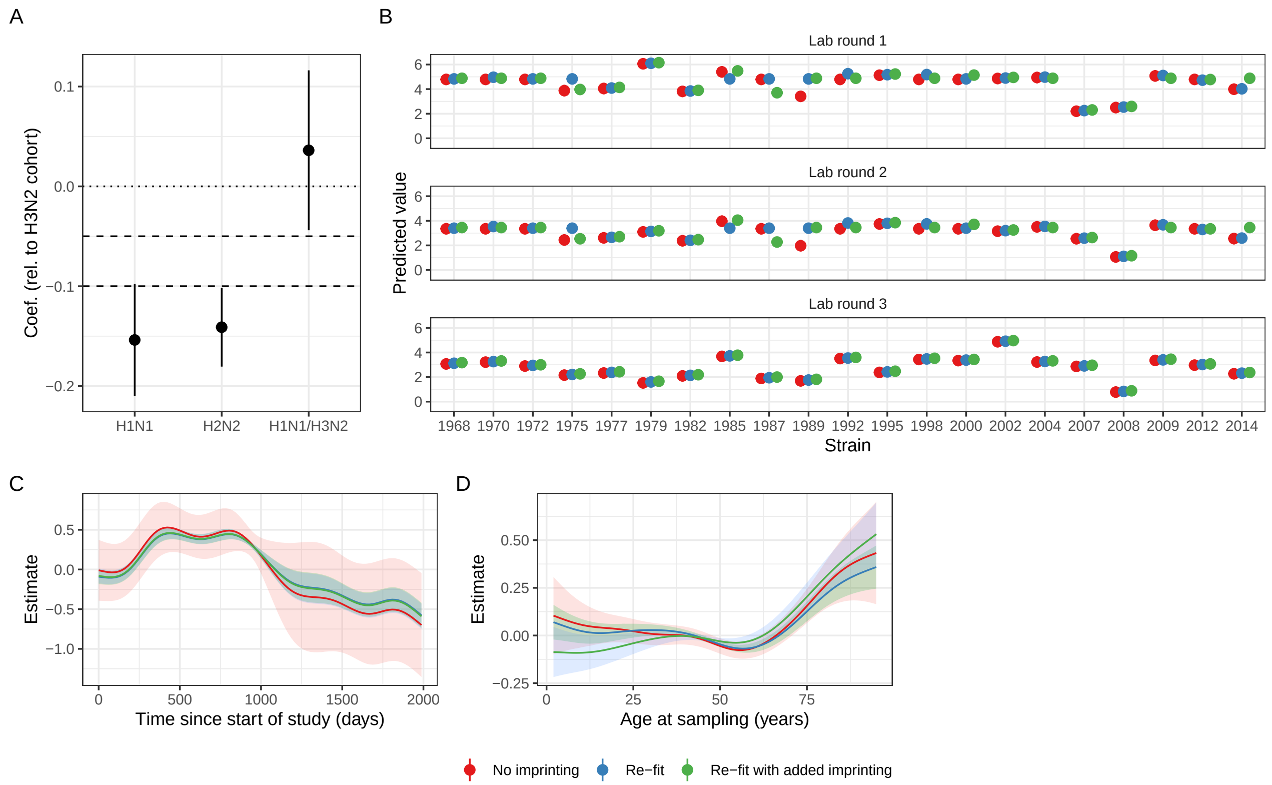

Figure S3: **Comparison of the coefficients and splines to check for parameter identifiability.** As in SI Fig S2, but with the inclusion of age at sampling.

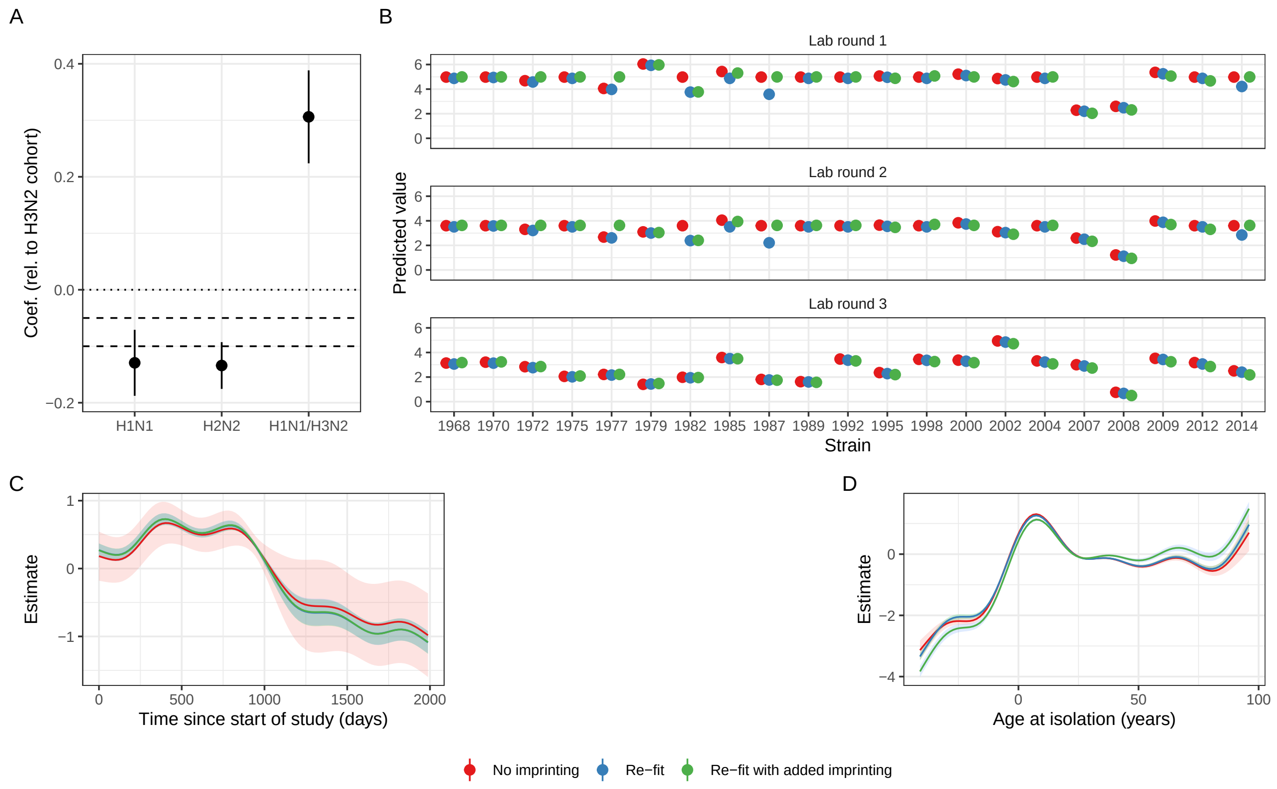

Figure S4: **Comparison of the coefficients and splines to check for parameter identifiability.** As in SI Fig S2, but with the inclusion of age at isolation.

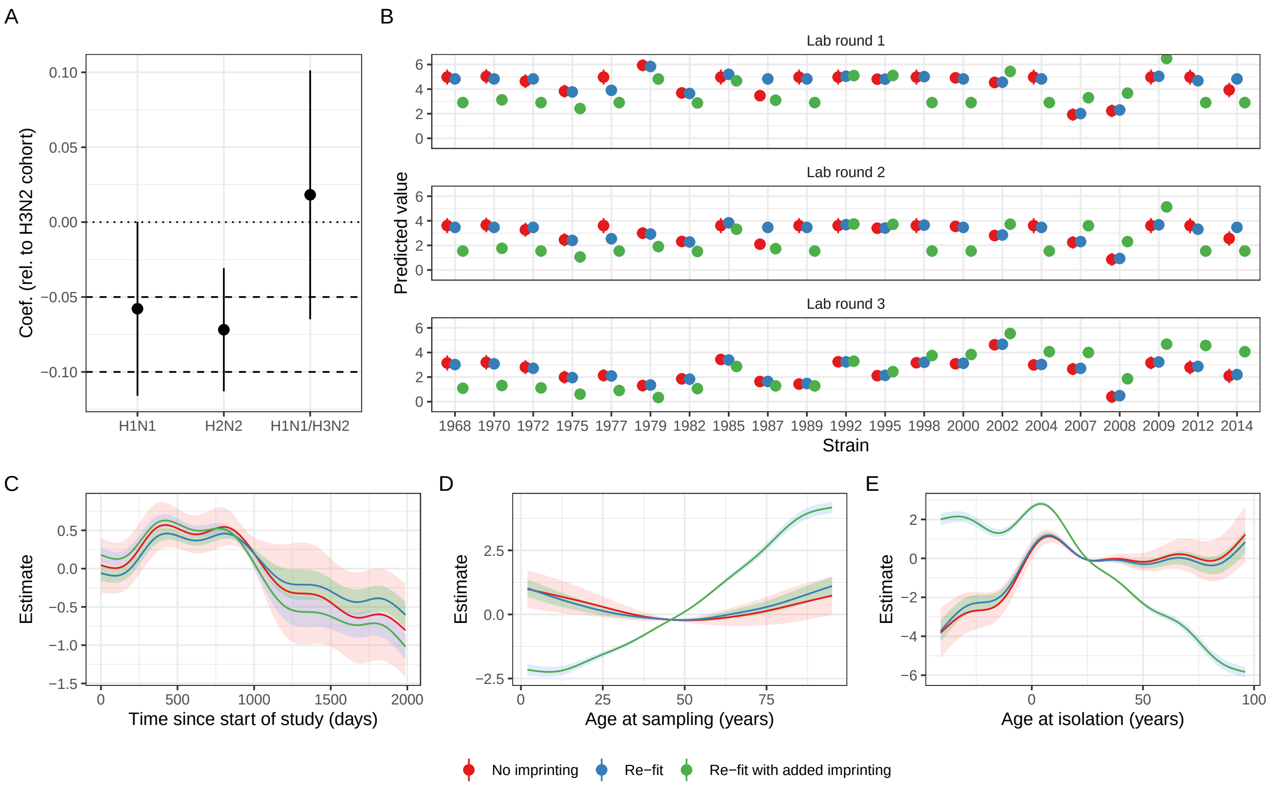

Figure S5: **Comparison of the coefficients and splines to check for parameter identifiability.** As in SI Fig S2, but with the inclusion of both age at sampling and age at isolation.

#### Potential impact of maternal antibodies

Allowing for the possible protection from maternal antibodies during the first few months of life, and that a sizeable proportion of participants were born in the latter half of a year, we also repeated all analyses by effectively adding one year to the year of birth of each participant when determining the subtype of first exposure (imprinting variable). For example, the subtype of first exposure for a participant born in 1967 would be the subtype present in 1968 (for the non-probabilistic scenario) or would be sampled from probabilities corresponding to 1968 (for the probabilistic case). This is equivalent to shifting the curves in Fig 4 in the main text one year to the left.

As in the main text, the best model by BIC includes both age variables, and similarly, only the imprinting variable coefficient for the H2N2 cohort is consistent with the imprinting hypothesis (SI Fig S6 and SI Table S4). However, more broadly, the patterns change little when the imprinting variable accounts for maternal antibodies and distribution of births within a year by shifting the introduction of new subtypes by a year (SI Fig S6).

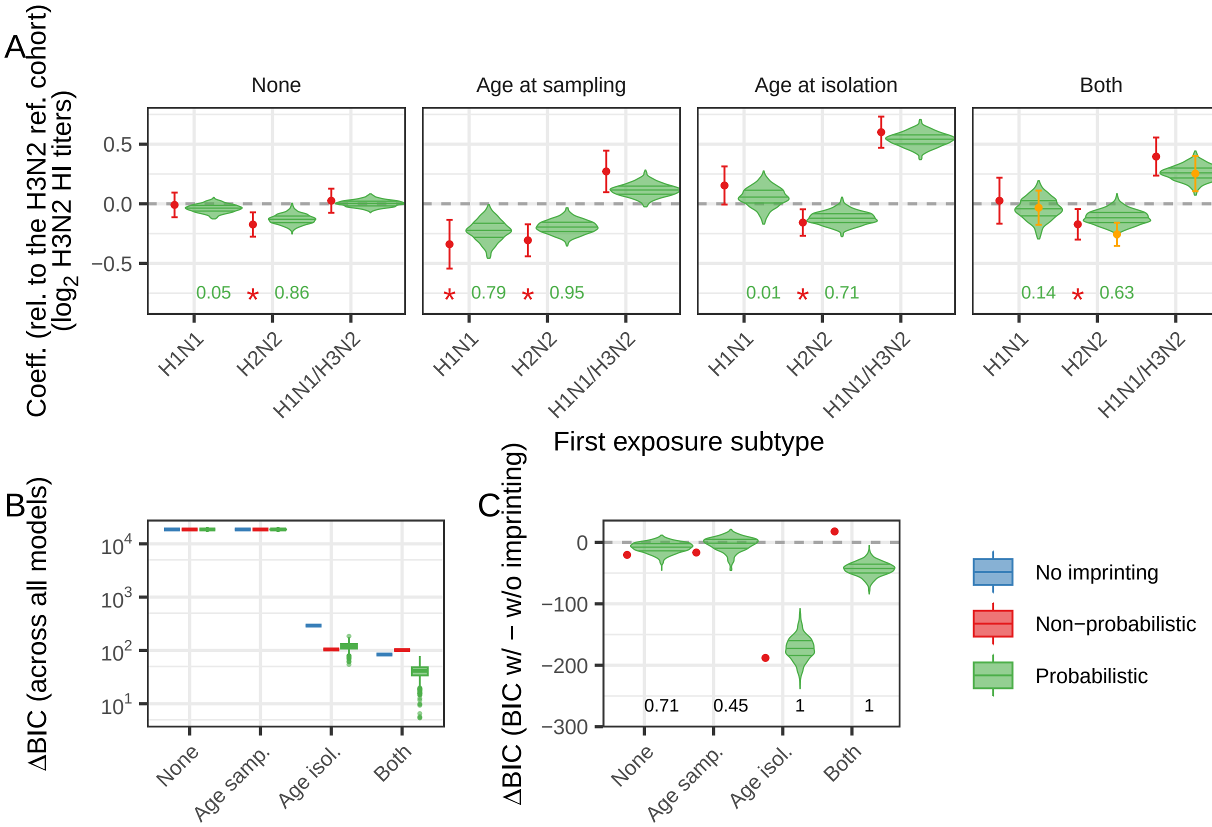

Figure S6: **Accounting for maternal antibodies.** Results analogous to those in Fig 1 in the main text, but in which we account for maternal antibodies and the fact that a significant proportion of participants are born in the second half of a year by effectively adding one to the year of birth, thus shifting the subtypes of first exposure.

#### Models fit to subsets of the data and simulation output

To further explore our findings, we repeat our analyses by focusing on specific subsets of the data. Since 1977, both H1N1 and H3N2 strains have co-circulated in the region. If infections in early life, rather than solely the first infection, are significant for imprinting, it is plausible that participants born on or after 1977 could exhibit systematic differences from those born prior to 1977 (i.e., having been imprinted by both H1N1 and H3N2). To explore this possibility, and given that we are primarily focussed on systematic differences in the titers of the earlier H1N1, H2N2, and H3N2 cohorts, we also fit models exclusively to data for individuals born before 1977.

Additionally, patterns in HI titers with respect to age at isolation or age at sampling differ qualitatively for the most recent strains (Fig 3 in the main text). Unlike HI titers against older strains, which exhibit a distinct peak for individuals aged around 5–10 years when the strain was circulating, this peak in titers is absent against the most recent strains. This discrepancy may be attributed to the considerably smaller proportion of individuals who were young when the more recent strains were circulating, as compared to older strains. To ensure that this qualitative difference did not introduce biases into our analyses, we also fit the models to HI titers against strains circulating prior to 2005.

When fitting the GAMs to participants born prior to 1977, the best model by BIC includes age at isolation, and coefficients are consistent with the imprinting hypothesis (SI Fig S7 and SI Table S3), but more broadly, results are generally not consistent with the imprinting hypothesis, and the inclusion of an imprinting variable does not improve model fit (middle row in SI Fig S7). On the other hand, the results when fitting the GAMs to strains that had circulated before 2005 were largely the same as those for the full dataset (bottom row in SI Fig S7); here too, the best model by BIC included both age variables and the probabilistic imprinting variable, but only the coefficient for the H2N2 cohort was consistent with the imprinting hypothesis.

The results for the models fit to subsets of the simulation output largely mirror the patterns in coefficients when fitting to subsets of the data (SI Fig S8).

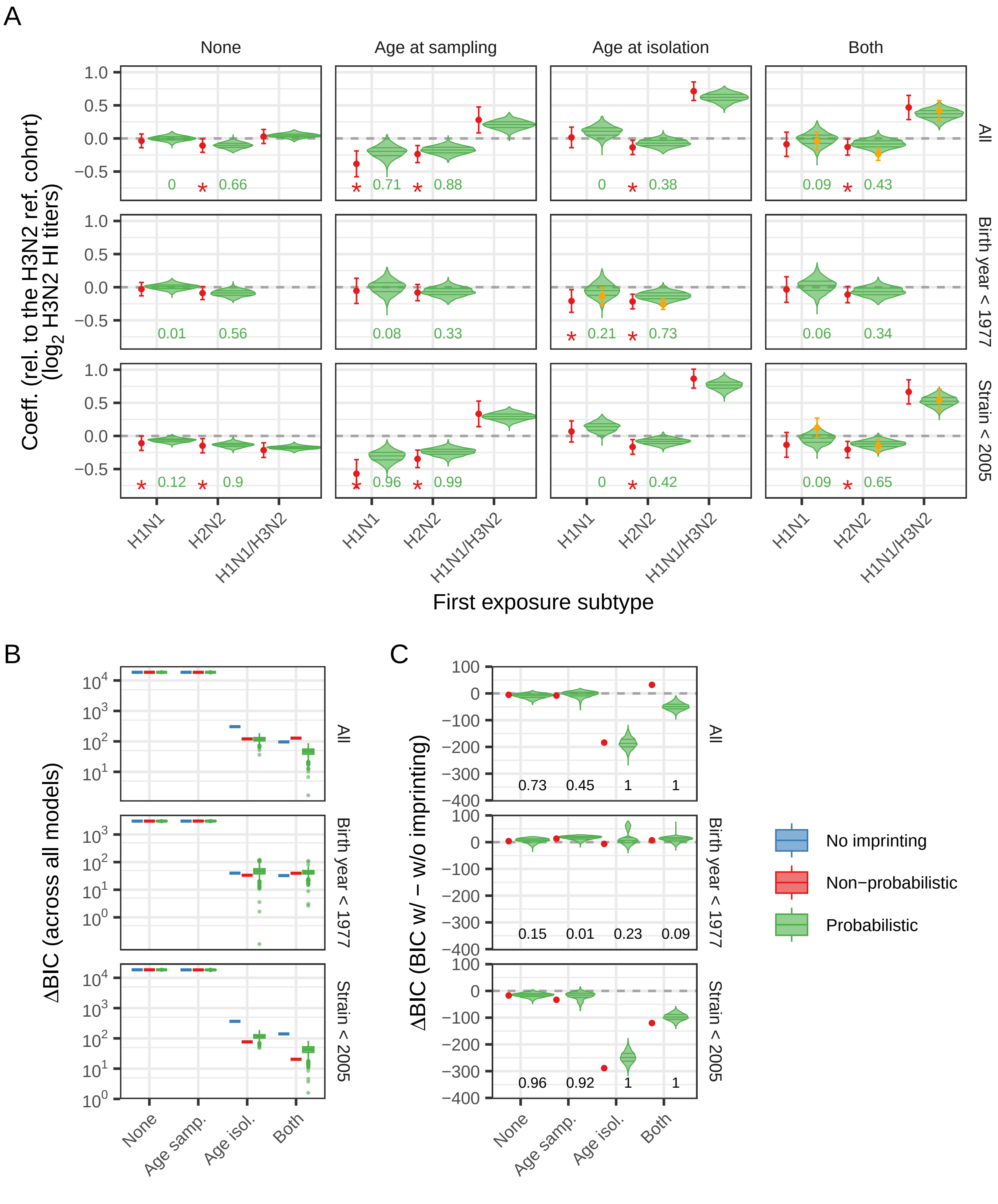

Figure S7: **Coefficients for models fit to subsets of the data.** Results analogous to those in Fig 1 in the main text, including results for fits to subsets of the data (rows of panels). The top row is the same as Fig 1 in the main text; the middle row are the results of the fits to data from participants born prior to 1977; the bottom row are the results of the fits to data for H3N2 strains that circulated prior to 2005.

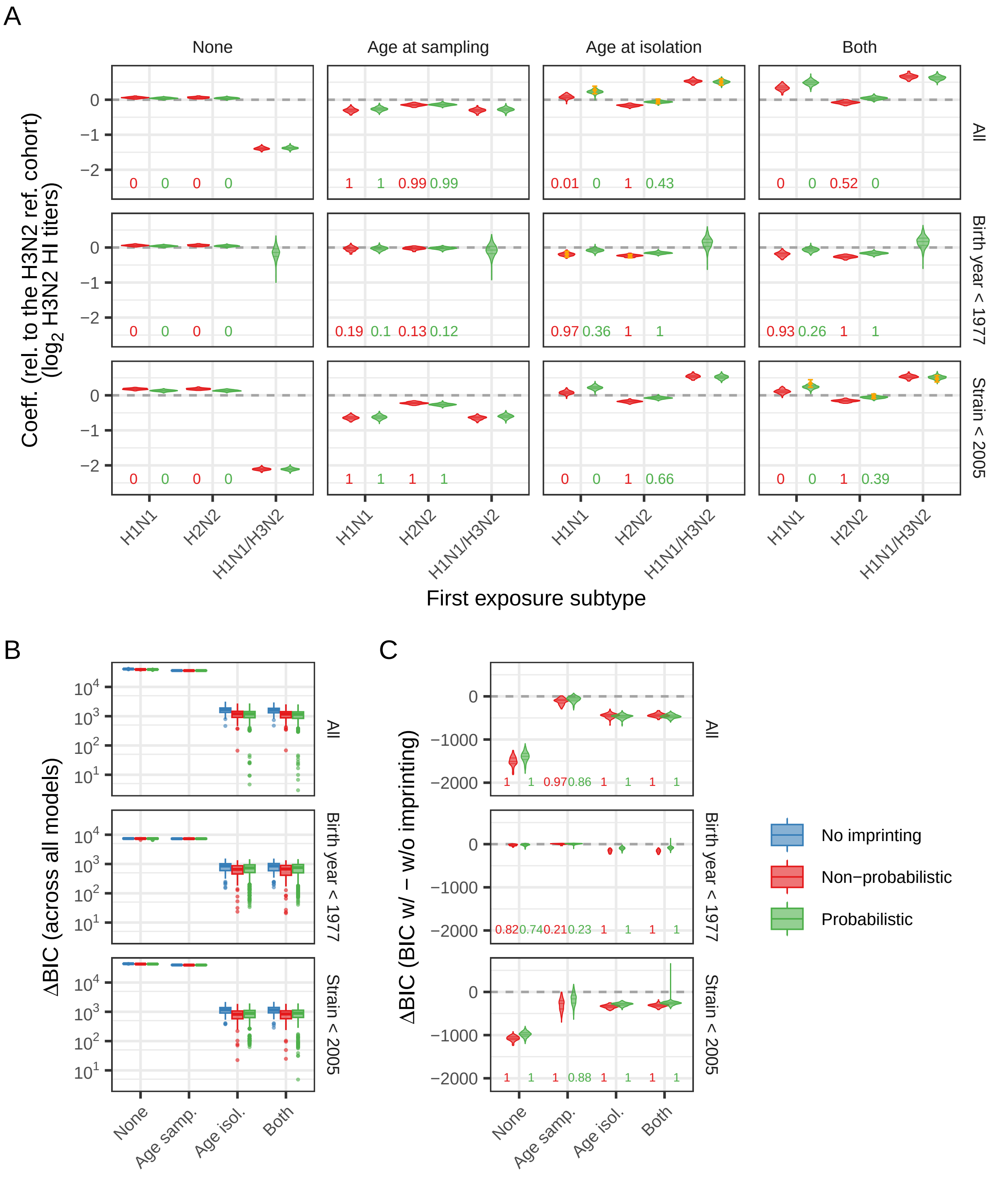

Figure S8: **Coefficients for models fit to subsets of the simulation output.** Results analogous to those in Fig 2 in the main text (for simulations without imprinting), including results for fits to subsets of the data (rows of panels). The top row is the same as the top row in Fig 2 in the main text; the middle row are the results of the fits to output from participants born prior to 1977; the bottom row are the results of the fits to output for H3N2 strains that circulated prior to 2005.

#### Randomising the demography for the models fit to the simulation output

We tested whether any of the patterns observed for the simulation output results were specific to the demography of the 777 participants. To do so, we generated a single dataset for 777 participants (without the 100 stochastic realisations) with an approximately uniform distribution of years of birth, by sampling across individuals and stochastic realisations of the null model output, and fit the GAM models (SI Fig S9).

Results in SI Fig S10 largely follow the same patterns as those without the randomisation of the demography of Fig 2 in the main text.

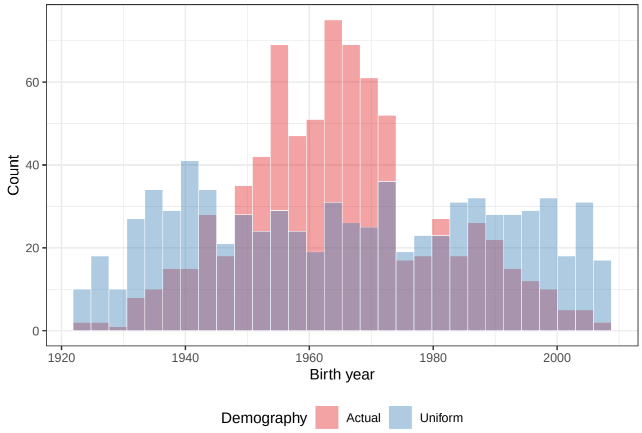

Figure S9: **Distribution of participants' years of birth.** Distribution of the years of birth across the 777 participants used in the null simulation models, and the uniform distribution obtained by resampling (see "Null simulation models") used in the robustness analysis.

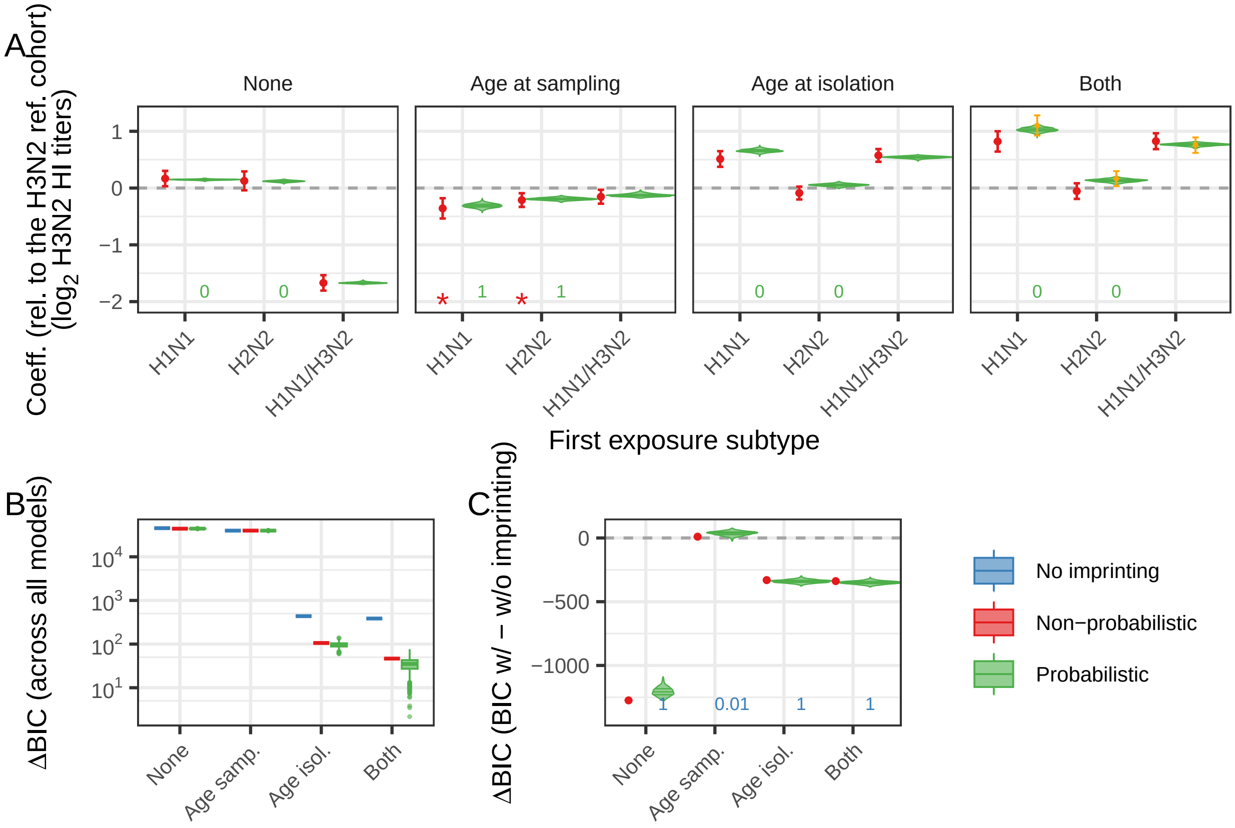

Figure S10: **Coefficients for a cohort with a uniform age distribution.** Results analogous to those in Fig 2 in the main text, but in which the the distribution of years of birth of the participants is uniform (see Fig S9). There are no significant changes in the results.

### Additional results

#### Best models

Table S3: **Best three models by BIC fit to H3N2 HI titers.** Best models are shown for the whole dataset ("All"), and for each of the subsets (sensitivity analyses).

| Age vars. | Coeff. | 95% CIs | Cohort | BIC | Dev. expl | RMSE |
|---|---|---|---|---|---|---|
| **All** | | | | | | |
| | -0.04 | (-0.174, 0.097) | H1N1 | | | |
| Both | -0.24 | (-0.331, -0.141) | H2N2 | 212632.1 | 0.60684 | 1.15948 |
| | 0.42 | (0.265, 0.572) | H1N1/H3N2 | | | |
| | 0.16 | (0.022, 0.304) | H1N1 | | | |
| Both | -0.14 | (-0.233, -0.048) | H2N2 | 212633.8 | 0.60689 | 1.15941 |
| | 0.38 | (0.226, 0.534) | H1N1/H3N2 | | | |
| | -0.16 | (-0.292, -0.023) | H1N1 | | | |
| Both | -0.20 | (-0.295, -0.115) | H2N2 | 212638.8 | 0.60688 | 1.15943 |
| | 0.53 | (0.384, 0.675) | H1N1/H3N2 | | | |
| **Birth year** < 1977 | | | | | | |
| Age isol. | -0.14 | (-0.269, -0.004) | H1N1 | 156670.0 | 0.58117 | 1.12337 |
| | -0.25 | (-0.335, -0.164) | H2N2 | | | |
| Age isol. | 0.14 | (-0.003, 0.276) | H1N1 | 156670.1 | 0.58145 | 1.12299 |
| | -0.18 | (-0.265, -0.088) | H2N2 | | | |
| Age isol. | -0.19 | (-0.327, -0.062) | H1N1 | 156671.6 | 0.58139 | 1.12308 |
| | -0.25 | (-0.331, -0.160) | H2N2 | | | |
| **Strain** < 2005 | | | | | | |
| | 0.12 | (-0.024, 0.270) | H1N1 | | | |
| Both | -0.15 | (-0.249, -0.057) | H2N2 | 162071.4 | 0.66304 | 1.07201 |
| | 0.55 | (0.388, 0.707) | H1N1/H3N2 | | | |
| | -0.01 | (-0.163, 0.137) | H1N1 | | | |
| Both | -0.13 | (-0.231, -0.024) | H2N2 | 162073.0 | 0.66296 | 1.07213 |
| | 0.66 | (0.509, 0.812) | H1N1/H3N2 | | | |
| | 0.02 | (-0.131, 0.162) | H1N1 | | | |
| Both | -0.15 | (-0.253, -0.044) | H2N2 | 162075.1 | 0.66296 | 1.07214 |
| | 0.63 | (0.472, 0.791) | H1N1/H3N2 | | | |

Table S4: **Best three models by BIC fit to H3N2 HI titers, while accounting for maternal antibodies.**

| Age vars. | Coeff. | 95% CIs | Cohort | BIC | Dev. expl | RMSE |
|---|---|---|---|---|---|---|
| | -0.03 | (-0.177, 0.111) | H1N1 | | | |
| Both | -0.26 | (-0.353, -0.159) | H2N2 | 212644.2 | 0.60672 | 1.15967 |
| | 0.25 | (0.106, 0.402) | H1N1/H3N2 | | | |
| | 0.02 | (-0.111, 0.149) | H1N1 | | | |
| Both | -0.17 | (-0.267, -0.083) | H2N2 | 212649.7 | 0.60665 | 1.15977 |
| | 0.34 | (0.182, 0.491) | H1N1/H3N2 | | | |
| | -0.27 | (-0.404, -0.130) | H1N1 | | | |
| Both | -0.29 | (-0.385, -0.196) | H2N2 | 212649.7 | 0.60674 | 1.15964 |
| | 0.38 | (0.226, 0.541) | H1N1/H3N2 | | | |

Table S5: **Best three models by BIC fit to H1N1 HI titers.**

| Age vars. | Coeff. | 95% CIs | Cohort | BIC | Dev. expl | RMSE |
|---|---|---|---|---|---|---|
| | -0.09 | (-0.423, 0.251) | H1N1 | | | |
| Age samp. | -0.31 | (-0.527, -0.085) | H2N2 | 25060.2 | 0.48913 | 1.37260 |
| | -0.09 | (-0.421, 0.232) | H1N1/H3N2 | | | |
| | 0.18 | (-0.149, 0.506) | H1N1 | | | |
| Age samp. | -0.18 | (-0.401, 0.035) | H2N2 | 25068.0 | 0.48957 | 1.37202 |
| | -0.14 | (-0.454, 0.172) | H1N1/H3N2 | | | |
| | 0.39 | (0.058, 0.714) | H1N1 | | | |
| Age samp. | -0.02 | (-0.232, 0.189) | H2N2 | 25069.2 | 0.48943 | 1.37220 |
| | -0.25 | (-0.545, 0.053) | H1N1/H3N2 | | | |

Table S6: **Best three models by BIC fit to simulation output.** Best models are shown for simulations without imprinting, and with an imprinting mechanism ("increased boosting"), as well as for subsets of the simulation output (without imprinting).

| Age vars. | Coeff. | 95% CIs | Cohort | BIC | Dev. expl | RMSE |
|---|---|---|---|---|---|---|
| **All (null model; no imprinting)** | | | | | | |
| | 0.28 | (0.173, 0.386) | H1N1 | | | |
| Age isol. | -0.05 | (-0.116, 0.019) | H2N2 | 84976.1 | 0.85284 | 0.80167 |
| | 0.51 | (0.417, 0.599) | H1N1/H3N2 | | | |
| | 0.47 | (0.347, 0.587) | H1N1 | | | |
| Both | 0.03 | (-0.044, 0.107) | H2N2 | 84979.1 | 0.85392 | 0.79873 |
| | 0.62 | (0.515, 0.725) | H1N1/H3N2 | | | |
| | 0.25 | (0.141, 0.355) | H1N1 | | | |
| Age isol. | -0.05 | (-0.120, 0.015) | H2N2 | 84980.8 | 0.85284 | 0.80167 |
| | 0.53 | (0.441, 0.625) | H1N1/H3N2 | | | |
| **All (positive control; imprinting via increased boosting)** | | | | | | |
| | -0.96 | (-1.101, -0.812) | H1N1 | | | |
| Both | -1.49 | (-1.581, -1.397) | H2N2 | 96356.2 | 0.83633 | 0.95136 |
| | 0.57 | (0.442, 0.695) | H1N1/H3N2 | | | |
| | -0.99 | (-1.128, -0.845) | H1N1 | | | |
| Both | -1.48 | (-1.566, -1.386) | H2N2 | 96401.9 | 0.83611 | 0.95310 |
| | 0.77 | (0.649, 0.897) | H1N1/H3N2 | | | |
| | -1.03 | (-1.176, -0.891) | H1N1 | | | |
| Both | -1.54 | (-1.636, -1.454) | H2N2 | 96456.3 | 0.83685 | 0.95348 |
| | 0.62 | (0.498, 0.748) | H1N1/H3N2 | | | |
| **Birth year** $<$ 1977 **(null model; no imprinting)** | | | | | | |
| Age isol. | -0.19 | (-0.278, -0.104) | H1N1 | 59083.1 | 0.85176 | 0.68874 |
| | -0.24 | (-0.296, -0.192) | H2N2 | | | |
| Both | -0.25 | (-0.347, -0.144) | H1N1 | 59104.0 | 0.85439 | 0.68495 |
| | -0.27 | (-0.328, -0.208) | H2N2 | | | |
| Both | -0.20 | (-0.301, -0.107) | H1N1 | 59105.8 | 0.85234 | 0.68741 |
| | -0.29 | (-0.344, -0.230) | H2N2 | | | |
| **Strain** $<$ 2005 **(null model; no imprinting)** | | | | | | |
| | 0.34 | (0.233, 0.445) | H1N1 | | | |
| Both | -0.03 | (-0.096, 0.038) | H2N2 | 49436.9 | 0.90249 | 0.56855 |
| | 0.47 | (0.382, 0.566) | H1N1/H3N2 | | | |
| | 0.36 | (0.261, 0.467) | H1N1 | | | |
| Both | -0.02 | (-0.085, 0.045) | H2N2 | 49441.7 | 0.90192 | 0.56780 |
| | 0.43 | (0.343, 0.524) | H1N1/H3N2 | | | |
| | 0.04 | (-0.050, 0.139) | H1N1 | | | |
| Age isol. | -0.20 | (-0.266, -0.142) | H2N2 | 49459.3 | 0.90187 | 0.56993 |
| | 0.53 | (0.447, 0.612) | H1N1/H3N2 | | | |

#### Model fits using deviance explained and RMSE

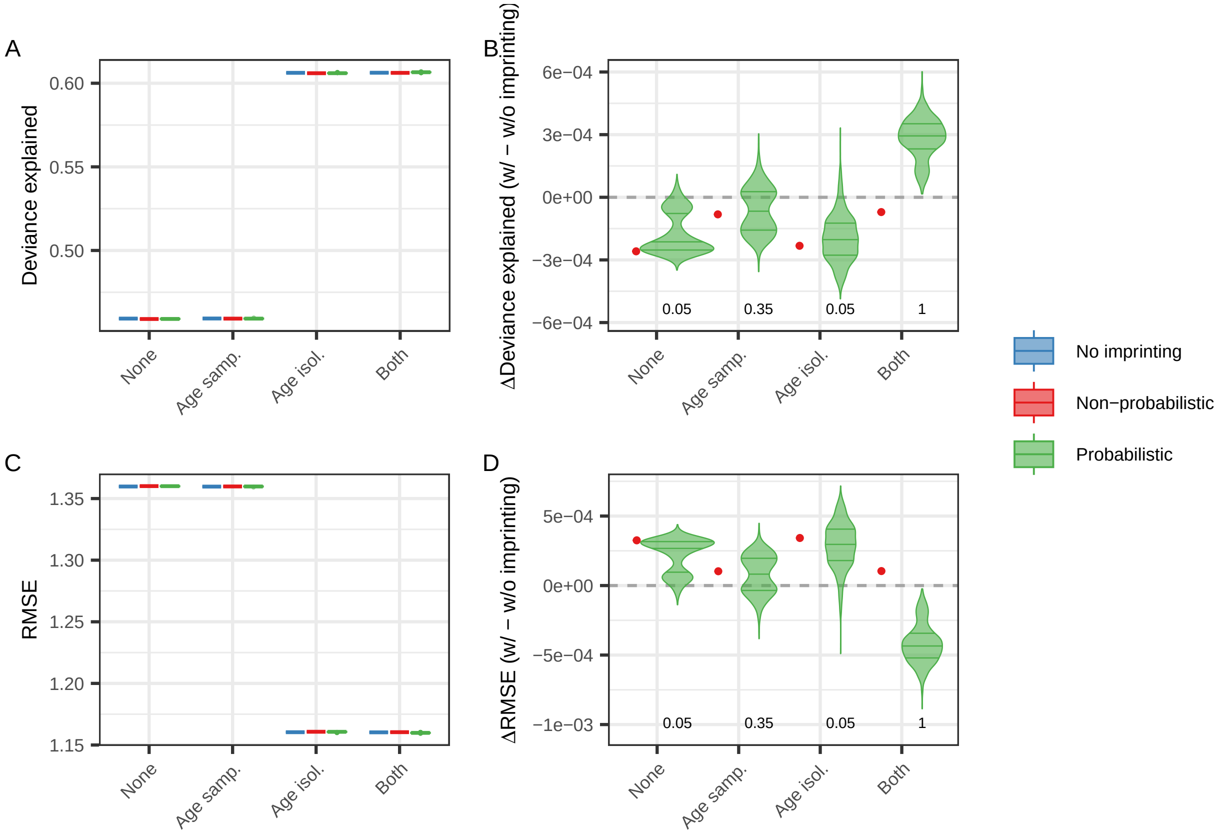

Figure S11: **Model fits using deviance explained and RMSE.** Results analogous to those in Fig 1B and C in the main text, but where model fits are described using deviance explained and root mean square error (RMSE) rather than BIC. Results in panels A and C reflect those in Fig 1B in the main text, with a significant improvement in model fit once age at isolation is included in the model. Panels B and D show that differences in model fit between models with and without the imprinting variable are small, and that by these metrics, the inclusion of the imprinting variable would often not be warranted, unless both age variables were included in the model.

#### Coefficients and splines for the models fit to data

Comparison of model splines and coefficients between models with and without the imprinting variable can reveal which model parameters might be absorbing any underlying birth cohort effects in absence of the imprinting variable, and therefore which parameters might still obscure any potential imprinting effects when the imprinting variable is included. When no age splines are included in the model, only the imprinting coefficient for the H2N2 cohort is significant, and there are few small shifts in the strain coefficients when comparing models with and without the imprinting variable (SI Fig S12), and a shift in the shape of the time since start of the study spline. On the other hand, when age at sampling is included, the imprinting coefficients are statistically significant. In absence of the imprinting variable, the imprinting pattern is absorbed by a shift in the age at sampling spline, compared to when the imprinting variable is included in the model (SI Fig S13). With the inclusion of age at isolation, again only the imprinting coefficient for the H2N2 cohort is significant, and the shifts in the age at isolation spline and strain coefficients are modest (SI Fig S14). However, it is with the inclusion of both age splines that we observe substantial shifts in the splines and coefficients, when comparing the model with the imprinting variable with the model without it (SI Fig S15). This latter result underscores how birth cohort patterns can potentially be traded off between multiple collinear variables, making it more challenging to interpret the imprinting variable.

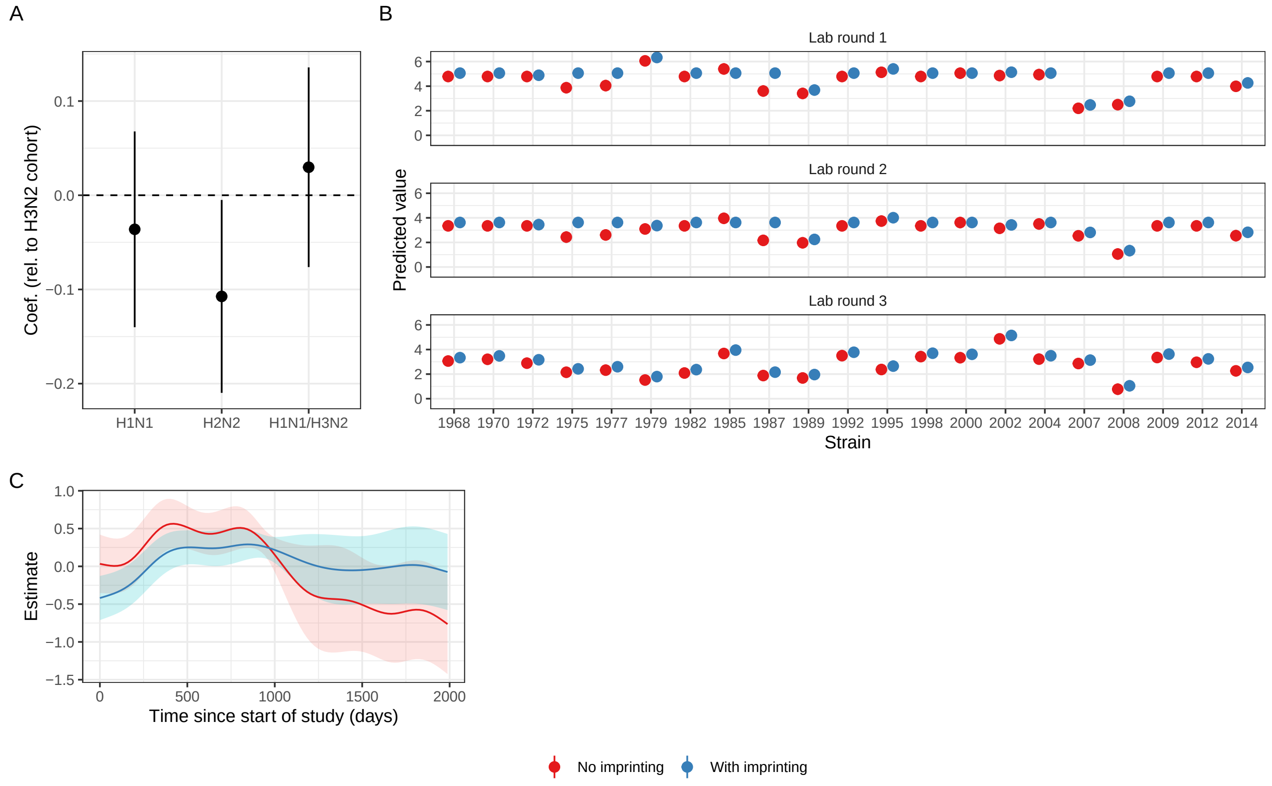

Figure S12: **Coefficients and splines for the models without an age spline.** These are shown with and without the non-probabilistic imprinting variable (corresponding to the models producing the red points in the first panel of Fig 1A in the main text).

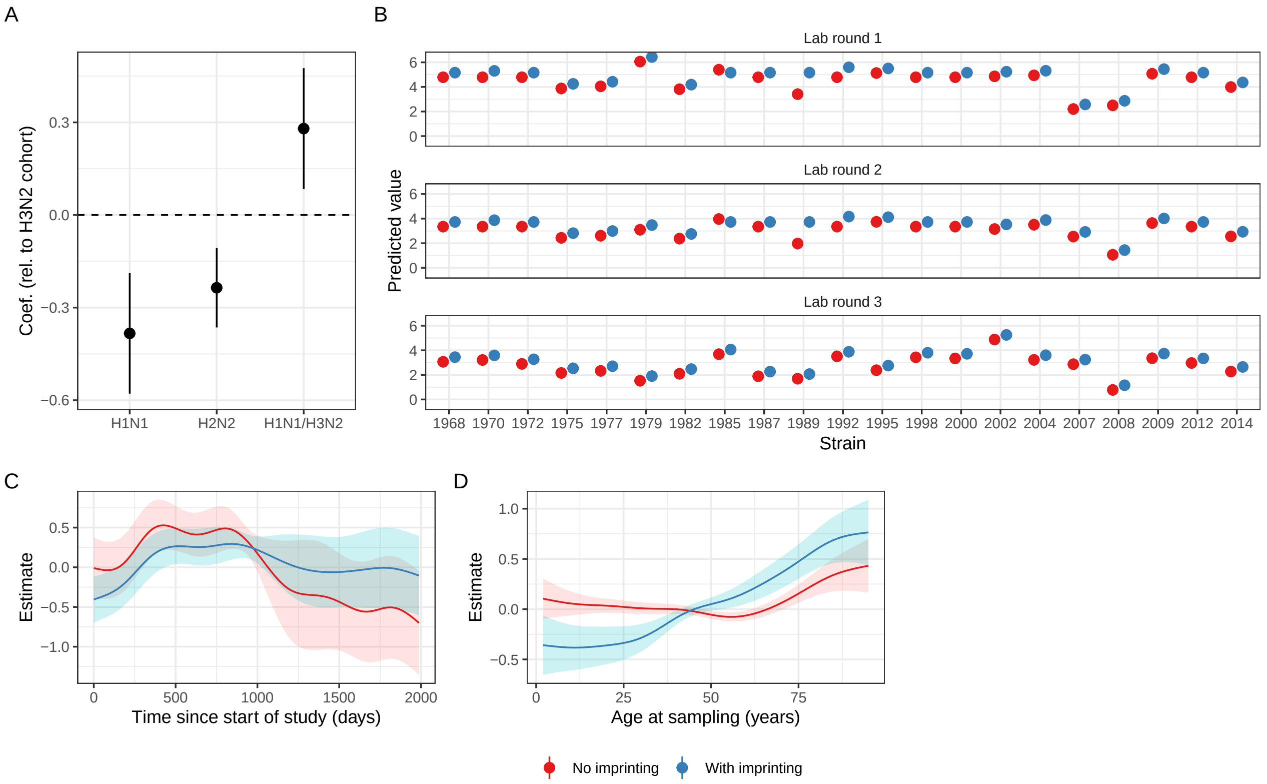

Figure S13: **Coefficients and splines for the models with a spline on age at sampling.** These are shown with and without the non-probabilistic imprinting variable (corresponding to the models producing the red points in the second panel of Fig 1A in the main text).

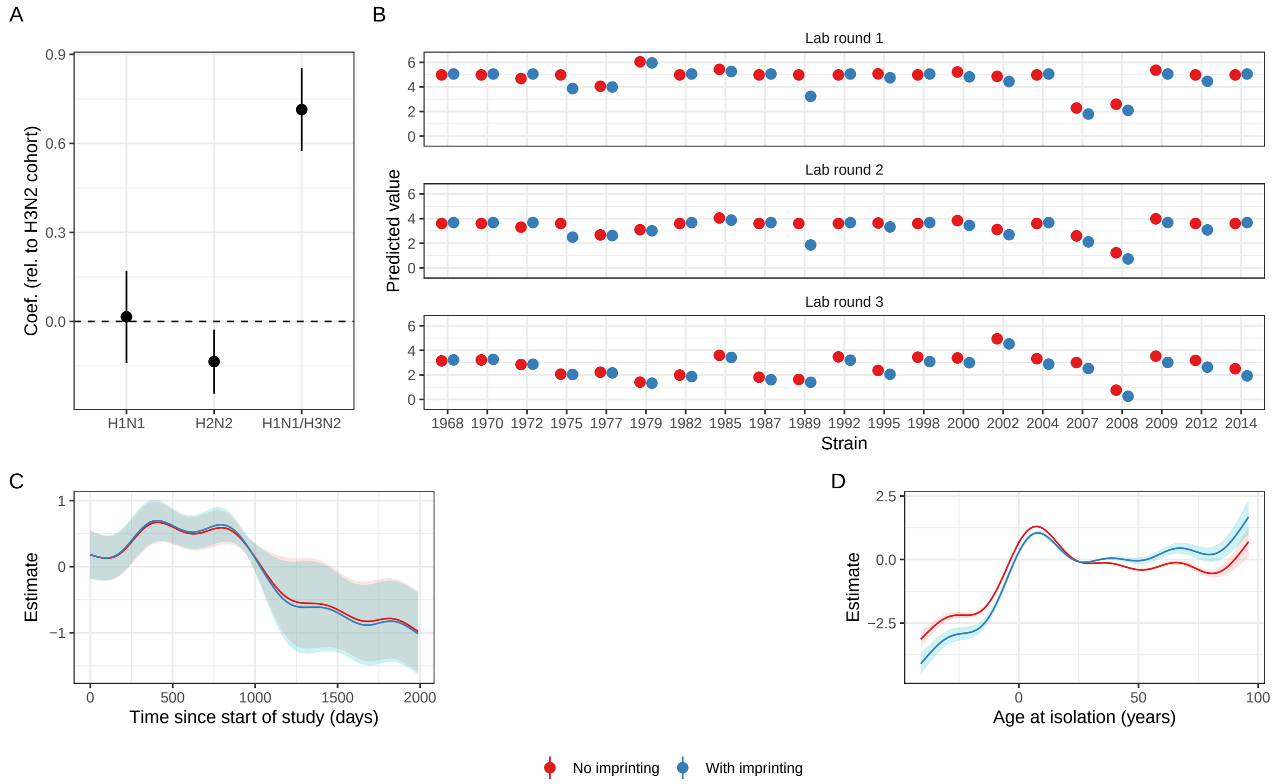

Figure S14: **Coefficients and splines for the models with a spline on age at isolation.** These are shown with and without the non-probabilistic imprinting variable (corresponding to the models producing the red points in the third panel of Fig 1A in the main text).

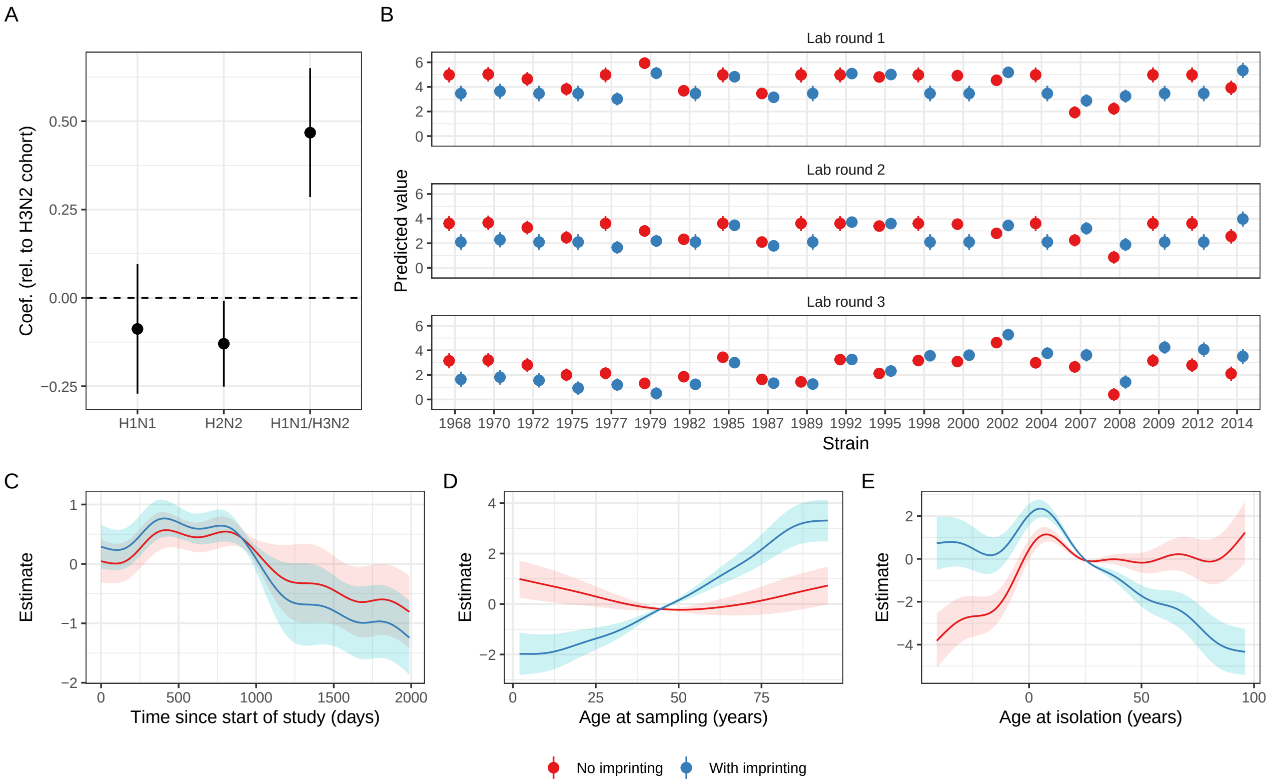

Figure S15: **Coefficients and splines for the models with splines on age at sampling and isolation.** These are shown with and without the non-probabilistic imprinting variable (corresponding to the models producing the red points in the third panel of Fig 1A in the main text).

#### Results for the models fit to H1N1 titers

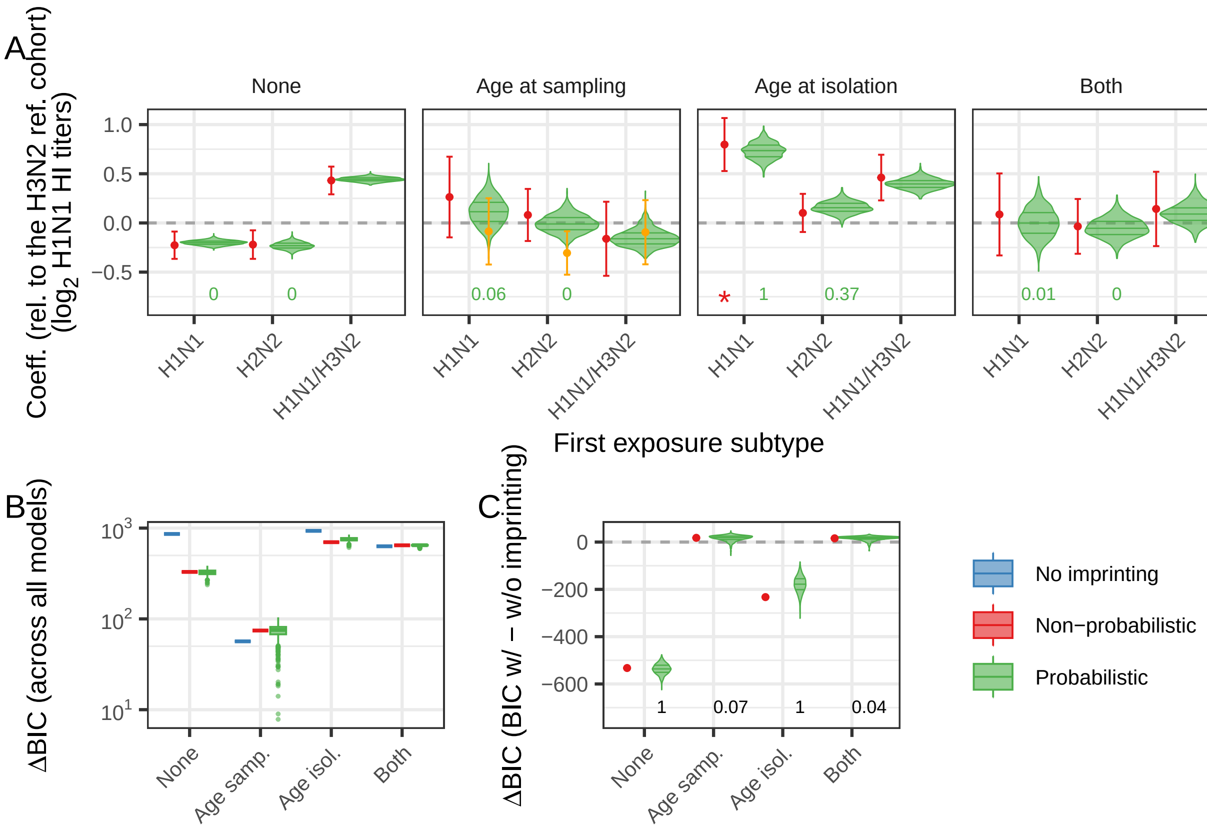

Figure S16: **Coefficients for models fit to H1N1 titers.** Results analogous to those in Fig 1 in the main text, but where models are fit to H1N1 titers. Here, under the imprinting hypothesis we would expect coefficients for the H1N1 and H2N2 cohorts to be systematically higher than those for the (reference) H3N2 cohort (i.e., we would expect positive coefficients for the two cohorts). As for Fig 1 in the main text, red stars highlight coefficients that align with the imprinting hypothesis, and the numbers indicate the proportion of the 1000 coefficients that are both statistically significant and consistent with imprinting. Orange points indicate the coefficients for the best model by BIC.
